# Efficacy and safety of TiNO-coated stents versus drug-eluting coronary stents. Systematic literature review and meta-analysis

**DOI:** 10.1101/2020.12.19.20248564

**Authors:** Frederic C. Daoud, Louis Létinier, Nicholas Moore, Pierre Coste, Pasi P. Karjalainen

## Abstract

**Objectives:** To compare clinical outcomes after percutaneous coronary intervention (PCI) using titanium-nitride-oxide coated stents (TiNOS) versus drug-eluting stents (DES) in coronary artery disease (CAD) including acute coronary syndrome (ACS).

**Design:** Prospective systematic literature (SLR) conducted according to PRISMA. Medline, Embase, Cochrane, Web of Science were searched in March 2018 and updated.

**Setting:** Interventional cardiology.

**Participants:** Patients with CAD, including ACS, requiring PCI.

**Interventions:** All prospective randomized controlled trials (RCTs) that compared clinical outcomes after PCI with DES versus TiNOS.

**Outcome measures:** The pooled risk ratios (RR), TiNOS over DES, with 95% confidence intervals (CI) are computed for device-oriented Major Adverse Cardiac Events (MACE), non-fatal myocardial infarction (MI), cardiac death (CD), clinically driven target lesion revascularization (TLR), probable or definite stent thrombosis (ST), total mortality, at one to five years after PCI. Pooled RRs are stratified according to baseline ACS versus other CAD. Sensitivity analysis (SA) and certainty of the evidence are rated per GRADE.

**Results:** Five RCTs are eligible with 1,855 patients with TiNOS versus 1,363 with DES at 1-year follow-up and 783 versus 771 at 5-year. Three RCTs included patients with ACS only. One-year RRs in ACS are: MACE 0.93 [0.72, 1.20], MI 0.48 [0.31, 0.73], CD 0.66 [0.33, 1.31], TLR 1.55 [1.10, 2.19] and ST 0.35 [0.20, 0.64]. One-year MACE, MI, and ST are robust to SA. The certainty of the evidence is high in MACE, moderate in MI, and low or very low in the other endpoints. There are too few observations to conclude about other CAD and 5-year outcomes. However, 5-year interim results are consistent with 1-year conclusions.

**Conclusions:** A similar risk of MACE is found in TiNOS and DES, with potentially fewer MI and ST but more TLR in TiNOS. TiNOS are safe and effective in ACS at 1-year follow-up.

**Registration:** PROSPERO CRD42018090622

**Strengths and limitations of this study:**

- Strengths:
  - The level of certainty of the evidence is high for the primary endpoint at one-year follow-up in patients treated for acute coronary syndrome.
  - The primary endpoint and critical secondary endpoints are robust to sensitivity analysis.

- Limitations:
  - Outcomes in patients treated for chronic coronary artery disease cannot be analyzed.
  - The level of certainty of the evidence of secondary endpoints is moderate or low.
  - Analysis of five-year outcomes is still at an interim stage.

## Objectives

Percutaneous coronary interventions (PCI) with drug-eluting stents (DES) is the standard of care in Coronary Artery Disease (CAD), including Acute Coronary Syndrome (ACS).^1-6^ mTOR inhibitors such as everolimus and paclitaxel have been the main drug types used on stents to inhibit post-stenting restenosis.^7-9^ Their side effect is the increased risk of stent thrombosis (ST), requiring prolonged Dual AntiPlatelet Therapy (DAPT) with its own risk of complications.^10,11^ Titanium-nitride-oxide coated coronary stents (TiNOS), also designated “bioactive stents” (BAS) have a pharmacologically inactive, non-absorbable coating. Preclinical data has shown less neointimal hyperplasia with TiNOS than with bare-metal stents (BMS).^12,13,14^

Several trials comparing TiNOS with DES have been conducted but no systematic review of that evidence has been published so far.

## Participants – Interventions - Outcome measures

The question was specified using the PICOS framework^15^: **<u>P</u>**atients presented CAD including ACS. **<u>I</u>**ntervention was PCI using TiNOS. **<u>C</u>**omparator was PCI using DES. **<u>O</u>**utcomes were the device-oriented Major Adverse Cardiac Events (MACE) and the components of that composiste i.e., Cardiac death (CD), recurrent myocardial infarction (MI), and clinically driven target lesion revascularization (TLR). Probable or definite ST, and all-cause mortality (“total death”: TD) were also analyzed. Outcomes were assessed at 1-year and 5-year follow-ups. **<u>S</u>**tudy methods were prospective randomized controlled clinical trials (RCTs).

## Methods

This systematic literature review (SLR) is the first to compare the efficacy and safety of TiNOS *versus* DES in CAD. It was designed and conducted according to methods described in the Cochrane Handbook with the use of “Grading of Recommendations Assessment, Development and Evaluation” (GRADE). The protocol was registered in PROSPERO (CRD4201809062) before initiation. It is reported according to “Preferred Reporting Items for Systematic Reviews and Meta-Analyses” (PRISMA).^16-18^

Data recording and the meta-analysis were conducted in RevMan 5.3 (Review Manager Version 5.3 software. Copenhagen, Denmark: The Nordic Cochrane Centre, 2014). The certainty of evidence according was rated with GRADEpro GDT software 2020 on-line version, (https://gradepro.org). Additional analyses were performed in STATA 16 (StataCorp LP, College Station, Tx, USA) using the metan and metaprop packages.

## Data sources

MEDLINE, EMBASE, the Cochrane Library, and Web of Science (WoS) electronic databases, were queried on March 8, 2018, using their search engines. The search terms were: *((bioactive OR (Titanium AND nitride AND oxide) OR TiNO OR TNO OR BAS) AND stent) AND (DES OR (drug AND eluting AND stent)) AND (RCT OR ((randomized OR randomised) AND controlled AND trial))*. No exclusion filter was applied related to language, country, year, or any other aspect. The websites of AHA, TCT, ESC, EuroPCR, and clinicaltrials.gov were also searched. The queries were updated on July 22, 2020, when all initially identified RCTs were published to retrieve new evidence if any, from RCTs meeting the PICOS specifications of this SLR.

The downloaded record files were imported and pooled and sifted in EndNote X8 (Clarivate Analytics, Philadelphia, PA, USA). One reference only was selected when duplicates were identified. When differerent references concerned the same study, their information was pooled using the citation of the most recent one. Full articles were reviewed for all references.

## Study selection and Data extraction

Two reviewers (FD and LL) performed independently the following steps: (1)Exhaustive reference screening, (2)Reference classification according to the inclusion and exclusion criteria, (3)Extraction of study methods, (4)Patient baseline data, (5)Treatment data and results of each eligible RCT, (6)Individual eligible RCT risk of bias rating, (7)Assessment of the certainty of the evidence for each outcome variable according to GRADE. The results were recorded in RevMan 5.3 file.

Differences were adjudicated by a third reviewer (NM). The similarity of the definitions of endpoints across the RCTs was discussed with one investigator (PK). The risk of bias of individual RCTs was rated according to the criteria proposed by the Cochrane Collaboration with operator blinding as a separate item.

The screened studies were included if they met the following criteria: First-hand clinical evidence with prospective inclusion; patients with CAD treated with coronary PCI; implantation of either TiNOS or DES after the random allocation of the stent type; target outcomes reported at 1-year and/or 5-year follow-up; the outcomes reported as the number of patients who were included along with the number or proportion of them who presented an event of interest. Studies were ruled out if any of the study selection criteria were not met or if IRB/ethics committee approval and patient informed consent were not explicitly required.

The term ACS referred to patients who presented at baseline with ST-elevated myocardial infarction (STEMI), or non-ST-elevated myocardial infarction (NSTEMI), or unstable angina pectoris.

The definitions of outcomes aimed in this SLR protocol are those defined by the Academic Research Consortium (ARC).^19^ Data extraction is stratified according to patient clinical presentation, i.e., ACS *versus* with other CAD.

## Statistical analysis

The two treatment arms are compared for each endpoint using the risk ratio (RR) defined as *((n patients with an event in TiNOS)/(n patients in TiNOS))/((n patients with an event in DES)/(n patients in DES))*. As a result, RR > 1 reflects a higher frequency of events in the TiNOS arm than in the DES arm and conversely. Outcomes are analyzed on an intention-to-treat (ITT) basis. The numerator is the number of patients presenting an event and counted in the treatment arm they were randomized to and the denominator is the sample size of the corresponding arm.

For each endpoint, individual study RRs are calculated with their 95% confidence interval (CI [,]) and their degree of heterogeneity is determined. If no significant heterogeneity is detected, the pooled RR is calculated with its CI using the M-H method with a fixed-effect model.^16,20,21^ In the case of significant heterogeneity, pooling uses the M-H method with a random-effects model. The pooled analysis is stratified according to baseline clinical presentation, ACS *versus* other CAD. One-year and 5-year outcomes are analyzed separately. Sensitivity analysis is performed by iteratively recalculating the pooled RR after removing one eligible RCT. As a result, the impact of each RCT on the pooled RR is estimated. It estimates the impact of differences in stent generations and the potential risk of indirection related to differences in outcome definitions.

## Patient and public involvement

No patient involved.

## Results

### Overall

Study identification, screening, and selection are described in the PRISMA flowchart (Figure 1). One hundred and eleven references were identified and nine publications with first-hand data about five RCTs are eligible for inclusion in the meta-analysis.

**Figure 1.**
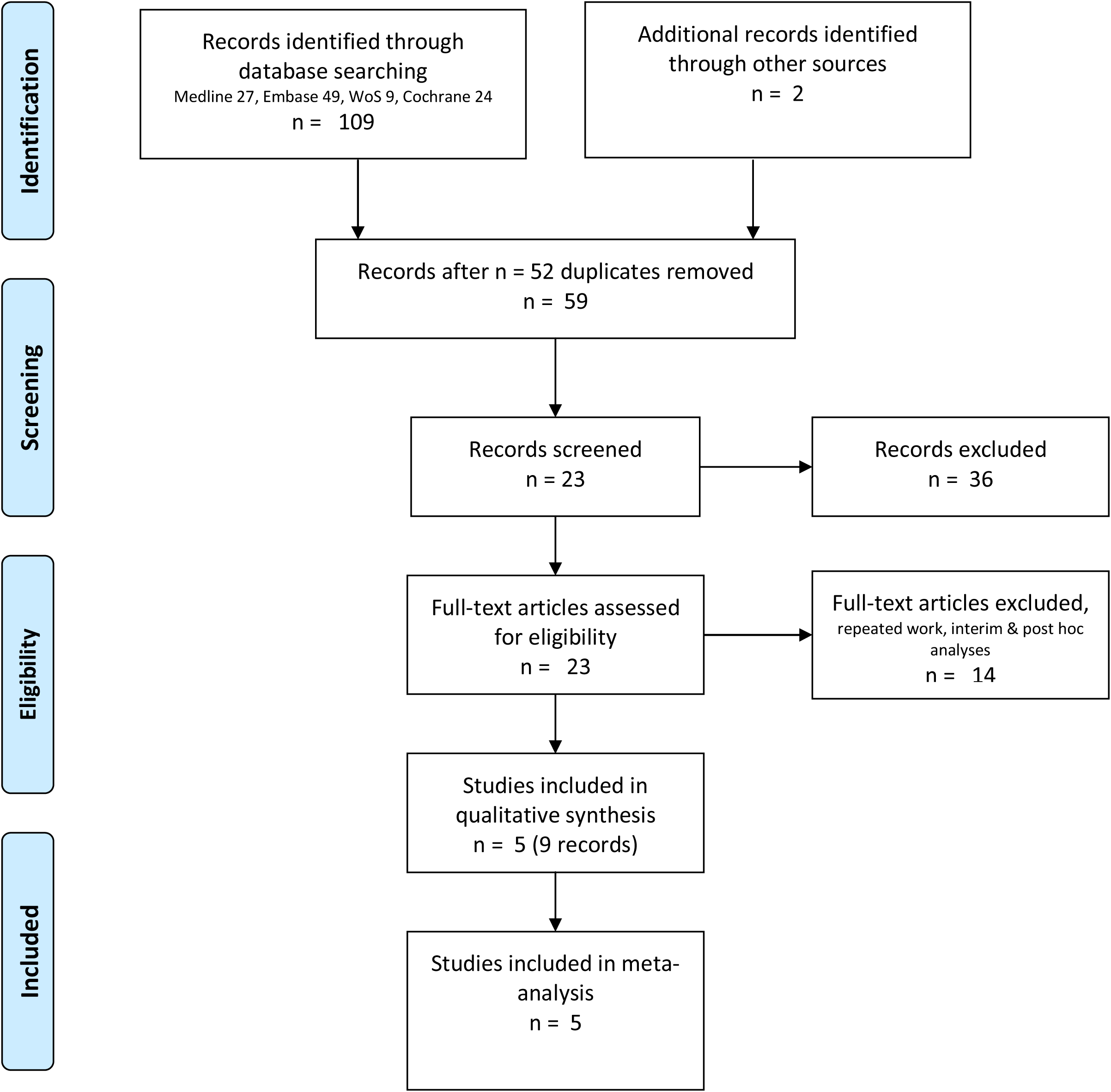
PRISMA flow chart.

The number of patients with 1-year follow-up data is 1,855 in the TiNOS arm *versus* 1,363 in the DES arm. Those numbers are 783 *versus* 773 patients at 5-year follow-up. Three RCTs enrolled only patients presenting with ACS and two enrolled and analyzed jointly patients presenting with ACS and other CAD. TIDE enrolled 143 patients with ACS (47%) and TITANIC-XV, 112 (64.7%). The baseline characteristics are summarized in Table 1. Funnel plots and Harbord tests did not detect a risk of publication bias concerning any of the endpoints at 1-year follow-up in all CAD (supplemental material).

**Table 1.** Baseline characteristics.

| Study | Age & prior events |  |  | Clinical presentation |  |  | Procedural data & medication |  |  | Attrition & cross-overs |
| --- | --- | --- | --- | --- | --- | --- | --- | --- | --- | --- |
| Stent |  | TiNOS | DES |  | TiNOS | DES |  | TiNOS | DES |  |
| TITAX-AMI<sup>22,23</sup><br>[NCT00495664] | Patients n<br>age<br>prior MI<br>prior PCI<br>prior CABG | 214<br>64±11<br>15%<br>10%<br>7% | 211<br>64±11<br>9%<br>5%<br>6% | NSTEMI<br>STEMI | 61%<br>39% | 54%<br>46% | stents in culprit<br>lesion<br>n<br>TSL (mm)<br>post-dilation<br>procedural success<br>DAPT 12-months | 1.1±0.3<br>18.5±6.4<br>42%<br>99.5%<br>31% | 1.1±0.4<br>19.2±7.2<br>35%<br>98.1%<br>65% | LFU<br>1y<br>TiNOS: 0<br>DES: 0<br>5y<br>TiNOS 3<br>DES: 7 |
| TIDE<sup>24</sup><br>[NCT00492908] | Patients n<br>age<br>prior MI<br>prior PCI<br>prior CABG | 152<br>65.9±9.0<br>27.6%<br>25.7%<br>7.9% | 150<br>63.4±10.5<br>28.7%<br>21.3%<br>25.3%<br>2.7% | Stable<br>CAD<br>NSTEMI<br>Unstable<br>angina | 57.9%<br>32.9%<br>9.2% | 47.3%<br>42.0%<br>10.7% | stents in culprit<br>lesion<br>n<br>TSL (mm)<br>device success<br>DAPT 12-months | 1.28±0.55<br>19.3±11.1<br>93.0% | 1.17±0.45<br>19.6±10.0<br>94.6% | LFU<br>1y<br>TiNOS: 0<br>DES: 2<br>5y : N.R. |
| TITANIC-XV<sup>25</sup><br>[NCT01510509] | Patients n<br>age<br>prior MI<br>prior PCI<br>prior CABG | 83<br>66.5±8.8<br>10.8%<br>8.4%<br>2.4% | 90<br>64.5±10.1<br>15.6%<br>11.1%<br>2.2% | NST ACS<br>Deducted<br>other CAD | 69.9%<br>30.1% | 60.0%<br>40.0% | stents in culprit<br>lesion<br>n<br>TSL (mm)<br>stent failure<br>DAPT 12 months | 1.1±0.3<br>18.72±8.20<br>N.R. | 1.1±0.3<br>21.63±9.65<br>N.R. | N.R. |
| BASE-ACS<sup>26,27</sup><br>[NCT00819923] | patients<br>age<br>prior MI<br>prior PCI<br>prior CABG | 417<br>63±12<br>13.4%<br>9.6%<br>4.8% | 410<br>63±12<br>9.8%<br>10.5%<br>4.1% | NSTEMI<br>STEMI<br>Unstable<br>angina | 49.4%<br>38.8%<br>11.8% | 45.6%<br>38.8%<br>15.6% | stents in culprit<br>lesion<br>n<br>TSL (mm)<br>post-dilation<br>stent failure<br>DAPT :<br>Aspirin : protocol<br>indefinite, actual<br>N.R.<br>Clopidogrel:<br>protocol 6 months,<br>actual N.R. | 1.15±0.38<br>20.8±9.4<br>42.2%<br>0.0% | 1.14±0.36<br>20.6±8.2<br>43.9%<br>1.0% | LFU<br>1y<br>TiNOS: 3<br>DES: 3<br>5y<br>TiNOS 29<br>DES: 28 |
| TIDES-ACS<sup>28,29</sup><br>[NCT02049229] | Patients n<br>age<br>prior MI<br>prior PCI<br>prior CABG | 989<br>62.7±10.<br>7.6%<br>7.0%<br>0.6% | 502<br>62.6±10.5<br>9.0%<br>6.6%<br>1.2% | NSTEMI<br>STEMI | 46.3%<br>44.9% | 45.0%<br>47.6% | stents in culprit<br>lesion<br>n<br>TSL (mm)<br>post-dilation<br>stent failure<br>DAPT 12-months | 1.13±0.38<br>20.5±7.8<br>33.0%<br>0.3%<br>80.3% | 1.14±0.37<br>20.6±7.2<br>38.0%<br>1.0%<br>86.0% | LFU 1y<br>TiNOS: 7<br>DES: 4 |

Four RCTs report or enable to deduct the number of cases of clinically driven TLR, CD, non-fatal recurrent MI, and the composite of CD or any non-fatal MI extended or clinically driven TLR. This results in a modified device-oriented MACE where MI “not clearly attributable to a nontarget vessel” is replaced by any MI. Given fatal MIs are counted as CD, this modification adds non-fatal MI in nontarget vessels.

In one RCT (TITANIC-XV), the primary endpoint is estimated from the available data as the sum of CD, any non-fatal MI, and any TLR assuming no overlap between those variables.

The risk of bias in individual RCTs is rated moderate or low except for the operator’s knowledge of the type of stent used during the intervention in all RCTs.

The pooled RRs of all outcomes at 1-year and 5-year follow-up are reported in Table 2 with CIs and sensitivity analyses. Results are reported overall and in the ACS subgroup. Given the 5-year follow-up of patients in the TIDES-ACS trial is ongoing, pooled RRs for 5-year outcomes are interim results.

**Table 2.** Pooled outcomes with sensitivity analysis.

|  | M-H fixed effects RR & 95% CI after the removal of: |  |  |  |  |  |
| --- | --- | --- | --- | --- | --- | --- |
| Endpoint | None: all RCTs included | TITAX-AMI | TIDE | TITANIC-XV | BASE-ACS | TIDES-ACS |
| **ALL CAD: ACS & Other CAD – 1 year follow-up** |  |  |  |  |  |  |
| MACE | 1.01 [0.81, 1.26] | 1.06 [0.83, 1.36] | 0.96 [0.75, 1.23] | 0.98 [0.78, 1.24] | 0.99 [0.76, 1.29] | 1.07 [0.82, 1.40] |
| CD or MI | 0.55 [0.39, 0.77] | 0.57 [0.39, 0.82] | 0.52 [0.36, 0.74] | 0.55 [0.39, 0.77] | 0.53 [0.36, 0.79] | 0.60 [0.40, 0.91] |
| Non-fatal MI | 0.52 [0.36, 0.76] | 0.51 [0.33, 0.78] | 0.48 [0.31, 0.73] | 0.52 [0.35, 0.76] | 0.60 [0.38, 0.93] | 0.51 [0.32, 0.82] |
| CD | 0.66 [0.33, 1.31] | 0.77 [0.37, 1.61] | 0.66 [0.33, 1.31] | 0.66 [0.33, 1.31] | 0.30 [0.11, 0.81] | 1.11 [0.43, 2.85] |
| Clinically driven TLR | 1.62 [1.20, 2.18] | 1.56 [1.13, 2.15] | 1.60 [1.15, 2.24] | 1.58 [1.16, 2.14] | 1.74 [1.22, 2.47] | 1.63 [1.14, 2.33] |
| Probable or definite ST | 0.39 [0.22, 0.69] | 0.46 [0.25, 0.84] | 0.35 [0.20, 0.64] | 0.39 [0.22, 0.69] | 0.36 [0.18, 0.72] | 0.38 [0.16, 0.87] |
| Definite ST | 0.51 [0.26, 0.99] | 0.49 [0.25, 0.97] | 0.46 [0.23, 0.92] | 0.51 [0.26, 0.99] | 0.62 [0.29, 1.36] | 0.52 [0.18, 1.44] |
| TD | 0.78 [0.48, 1.27] | 0.77 [0.46, 1.32] | 0.78 [0.47, 1.27] | 0.78 [0.48, 1.27] | 0.49 [0.26, 0.95]<sup>b</sup> | 1.22 [0.65, 2.28] |
| **ACS only – 1 year follow-up** |  |  |  |  |  |  |
| MACE | 0.93 [0.72, 1.20] | 0.97 [0.73, 1.30] | N.A. | N.A. | 0.86 [0.63, 1.19] | 0.95 [0.68, 1.33] |
| CD or MI | 0.52 [0.36, 0.75] | 0.53 [0.35, 0.80] | N.A. | N.A. | 0.48 [0.30, 0.75] | 0.56 [0.35, 0.90] |
| Non-fatal MI | 0.48 [0.31, 0.73] | 0.45 [0.27, 0.74] | N.A. | N.A. | 0.55 [0.33, 0.92] | 0.44 [0.25, 0.77] |
| CD | 0.66 [0.33, 1.31] | 0.77 [0.37, 1.61] | N.A. | N.A. | 0.30 [0.11, 0.81] | 1.11 [0.43, 2.85] |
| Clinically driven TLR | 1.55 [1.10, 2.19] | 1.46 [0.99, 2.15] | N.A. | N.A. | 1.69 [1.09, 2.63] | 1.53 [0.97, 2.40] |
| Probable or definite ST | 0.35 [0.20, 0.64] | 0.42 [0.22, 0.78] | N.A. | N.A. | 0.32 [0.15, 0.65] | 0.31 [0.12, 0.77] |
| Definite ST | 0.46 [0.23, 0.92] | 0.43 [0.21, 0.89] | N.A. | N.A. | 0.54 [0.24, 1.24] | 0.39 [0.12, 1.25] |
| TD | 0.78 [0.47, 1.27] | 0.77 [0.45, 1.32] | N.A. | N.A. | 0.47 [0.24, 0.93]<sup>b</sup> | 1.23 [0.64, 2.35] |
| **ALL CAD: ACS & Other CAD – 5 year follow-up - Interim Results** |  |  |  |  |  |  |
| MACE | 0.82 [0.66, 1.02] | 0.91 [0.70, 1.19] | 0.74 [0.58, 0.95]<sup>b</sup> | N.A. | 0.83 [0.62, 1.10] | Expected |
| CD or MI | 0.57 [0.42, 0.77] | 0.67 [0.46, 0.97] | 0.53 [0.38, 0.73] | N.A. | 0.53 [0.35, 0.80] | Expected |
| Non-fatal MI | 0.56 [0.39, 0.80] | 0.59 [0.37, 0.92] | 0.54 [0.37, 0.80] | N.A. | 0.57 [0.35, 0.93] | Expected |
| CD | 0.68 [0.39, 1.19] | 0.93 [0.47, 1.82] | 0.59 [0.31, 1.11] | N.A. | 0.55 [0.25, 1.24] | Expected |
| Clinically driven TLR | 1.01 [0.75, 1.35] | 1.00 [0.71, 1.42] | 0.94 [0.66, 1.32] | N.A. | 1.13 [0.77, 1.66] | Expected |
| Probable or definite ST | 0.30 [0.14, 0.61] | 0.45 [0.19, 1.05] | 0.25 [0.12, 0.55] | N.A. | 0.22 [0.07, 0.70] | Expected |
| Definite ST | 0.25 [0.11, 0.55] | 0.37 [0.14, 0.99] | 0.20 [0.09, 0.49] | N.A. | 0.22 [0.07, 0.70] | Expected |
| TD | 1.03 [0.74, 1.45] | 1.14 [0.75, 1.73] | 0.95 [0.65, 1.37] | N.A. | 1.05 [0.65, 1.69] | Expected |
| **ACS only – 5 year follow-up - Interim Results** |  |  |  |  |  |  |
| MACE<sup>a</sup> | 0.74 [0.58, 0.95] | 0.81 [0.59, 1.12]<sup>c</sup> | N.A. | N.A. | 0.65 [0.44, 0.95]<sup>c</sup> | Expected |
| CD or MI | 0.53 [0.38, 0.73] | 0.61 [0.40, 0.95] | N.A. | N.A. | 0.42 [0.25, 0.71] | Expected |
| Non-fatal MI | 0.54 [0.37, 0.80] | 0.56 [0.33, 0.94]<sup>c</sup> | N.A. | N.A. | 0.53 [0.30, 0.94]<sup>c</sup> | Expected |
| CD | 0.59 [0.31, 1.11] | 0.83 [0.38, 1.84]<sup>c</sup> | N.A. | N.A. | 0.33 [0.11, 1.00]<sup>c</sup> | Expected |
| Clinically driven TLR | 0.94 [0.66, 1.32] | 0.88 [0.56, 1.37]<sup>c</sup> | N.A. | N.A. | 1.03 [0.60, 1.76]<sup>c</sup> | Expected |
| Probable or definite ST<sup>a</sup> | 0.25 [0.12, 0.55] | 0.37 [0.15, 0.93]<sup>c</sup> | N.A. | N.A. | 0.13 [0.03, 0.57]<sup>c</sup> | Expected |
| Definite ST<sup>a</sup> | 0.20 [0.09, 0.49] | 0.28 [0.09, 0.85]<sup>c</sup> | N.A. | N.A. | 0.13 [0.03, 0.57]<sup>c</sup> | Expected |
| TD | 0.95 [0.65, 1.37] | 1.02 [0.63, 1.65]<sup>c</sup> | N.A. | N.A. | 0.85 [0.48, 1.53]<sup>c</sup> | Expected |

<sup>a</sup>sensitivity analysis in ACS at 5-year follow-up results in the RR and confidence intervals of individual RCTs; <sup>b</sup>borderline shift with 3 or fewer RCTs contributing to the pooled estimate; <sup>c</sup>results based on a single-trial; N.A. Not applicable

The stratified (ACS relative to other CAD) pooled RRs of effectiveness endpoints show no significant difference in 1-year MACE and a significantly higher rate of TLR with TiNOS (Figure 2). The stratified stratified pooled RRs of safety endpoints show no significant difference in 1-year CD, but significantly lower rates in non-fatal, MI and ST with TiNOS compared to DES (Figure 3). The pooled RR of TD is also not significant (supplemental file).

**Figure 2.**
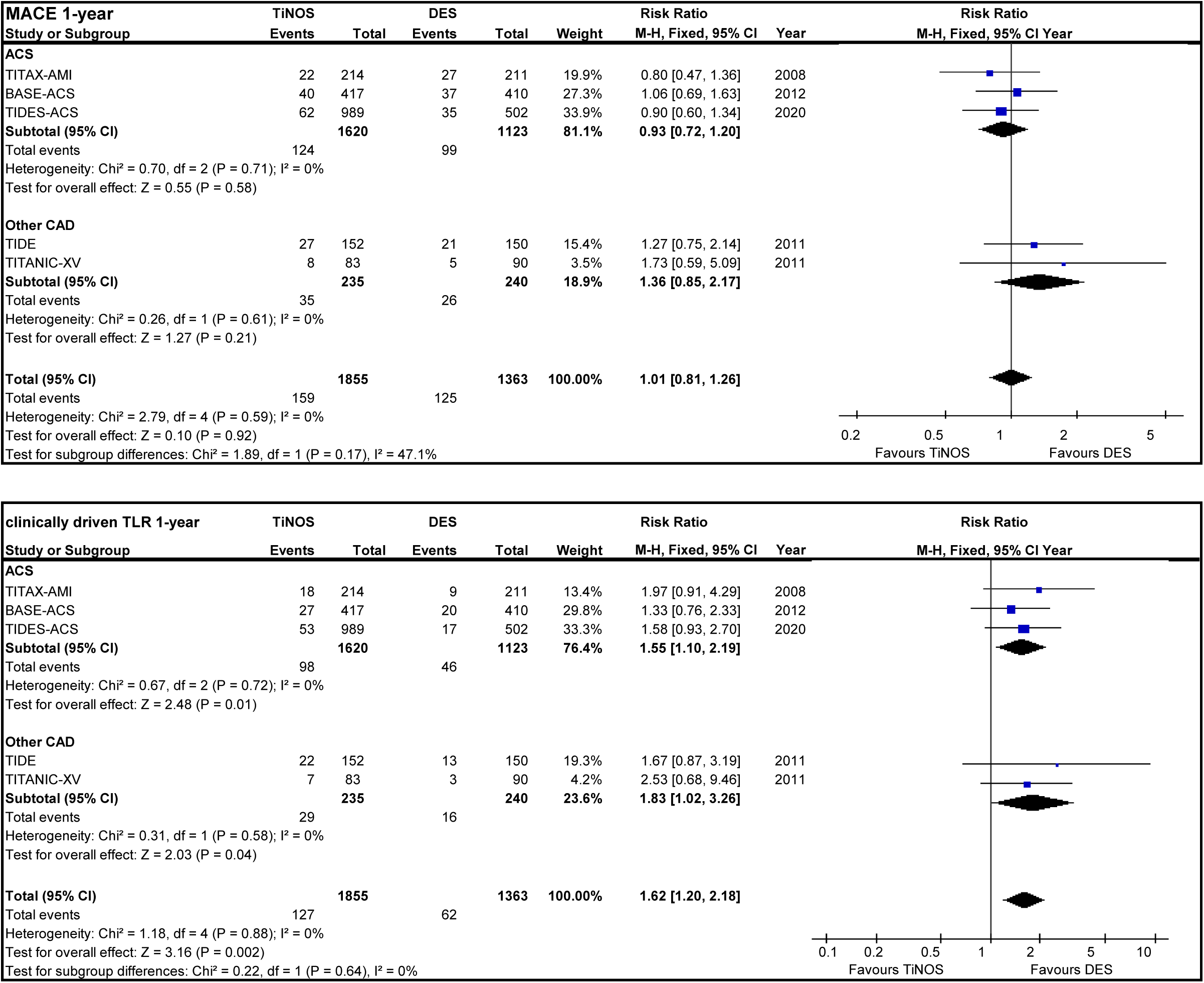
Effectiveness endpoints.

**Figure 3.**
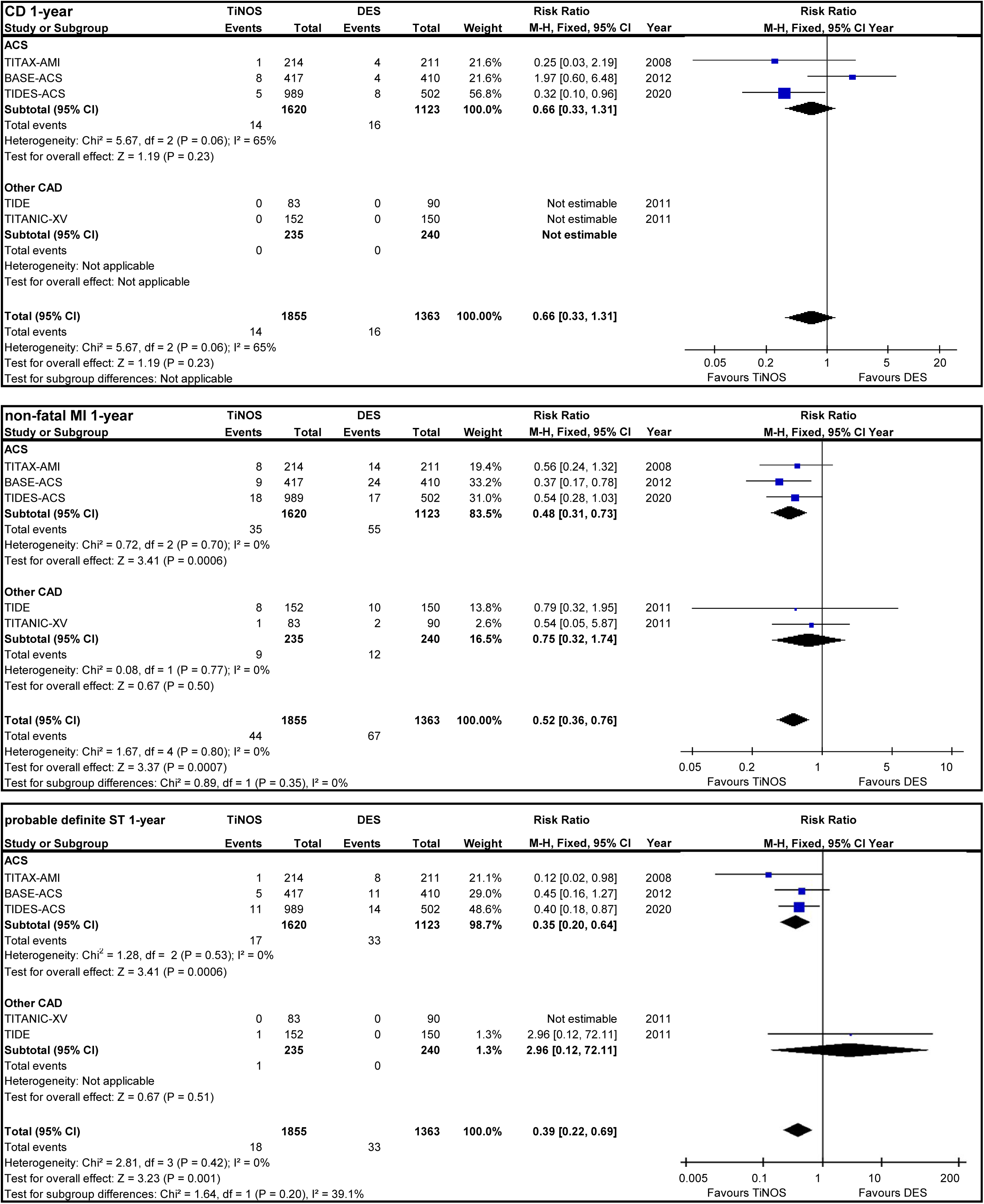
Safety endpoints.

The stratified pooled RRs analysis shows a much larger number of patients in the ACS subgroup compared to the other CAD subgroup (85.2% *versus* 14.8%). The overall effects are driven by the ACS subgroup. The results of the other CAD subgroup (TIDE and TITANIC-XV) are not interpretable because outcomes in patients with ACS are not reported separately. Therefore, the subsequent steps of this SLR are refocused on RCT about ACS only.

### ACS at 1-year follow-up

The pooled RRs of effectiveness and safety endpoints in MACE are consistent with overall results but heterogeneity, robustness to sensitivity analysis (Table 2) and the level of certainty of the underlying evidence according to GRADE (supplemental material) need to be assessed separately.

The RR of MACE presents no significant heterogeneity, is robust to sensitivity analysis, and the certainty of evidence is rated high. The RR of clinically driven TLR presents no significant heterogeneity but results are not robust to sensitivity analysis, and the certainty of the evidence is rated low.

The pooled RRs of CD and TD present significant heterogeneity, are not robust to sensitivity analysis, and the certainty of the evidence is rated very low.

The pooled RR of non-fatal MI presents no significant heterogeneity, is robust to sensitivity analysis, and the certainty of the evidence is rated moderate.

The pooled RR of probable or definite SE presents no significant heterogeneity, is robust to sensitivity analysis, and the certainty of the evidence is rated low.

## Discussion

TIDE and TITANIC-XV enrolled 255 patients with ACS but their data are not reported separately from the other patients. The impact of not including them in the ACS subgroup is limited given they represent 7.9% of all patients with ACS. The modified definition of device-oriented MACE results in including non-fatal MIs from nontarget vessels. One can reasonably assume the index stent does not affect those events. Ninety non-fatal MIs are reported. Assuming half of them are related to nontarget vessels (i.e., 45 cases), proportionality with sample size would lead to 27 fewer cases with TiNOS and 18 with DES, which would result in an RR of 0.19 [0.09, 0.42]. The inclusion of nontarget MIs thus results in a dilution that is favorable to DES. If the same numbers cases were removed from the count of MACE, the RR would be 0.98 [0.73, 1.30], which would not change the non-inferiority conclusion. The robustness to sensitivity analysis of the pooled RRs of MACE, MI, and ST in ACS, shows that the differences in DES generations, including the elution of paclitaxel and mTOR inhibitors do not significantly change those results. The interim 5-year pooled results will be updated when the final results of TIDES-ACS are published, but the current results are consistent with 1-year outcomes.

## Conclusions

This systematic literature review shows that titanium-nitride oxide coated-coronary stents and drug-eluting stents have a similar risk of device-oriented major adverse cardiac events at one-year follow-up in patients with an acute coronary syndrome. This result is robust and the level of certainty of the evidence is high. It also shows a lower risk of recurrent myocardial infarction and stent thrombosis with titanium-nitride oxide coated-coronary stents than with drug-eluting stents with a potentially higher risk of target lesion revascularization. Interim five-year pooled outcomes are consistent with one-year outcomes. These results show that the titanium-nitride oxide coated-coronary stents are safe and effective in acute coronary syndrome at one-year follow-up.

## Supporting information

PRISMA checklist

## Data Availability

This work used published summary data only.

## Supplemental material

- Details of methods applied in this SLR.
- Methods of individual RCTs
- Table of the number of patients with events – ITT
- Funnel plots and results of Harbord tests.
- Summary of risk of bias in individual RCTs
- Pooled RR of CD or MI, and TD
- GRADE certainty of evidence assessment
- PRISMA checklist

## Contributors

Frederic Daoud (FD): Main reviewer and meta-analyst with LL.

Louis Létinier (LL): Main reviewer and meta-analyst with with FD.

Nicholas Moore (NM): Third reviewer. Adjudicated disagreements.

Pierre Coste (PC): Advised on protocol and analysis.

Pasi Karjalainen (PK): Provided additional information about the methods of the individual trials.

## Data sharing statement

All data included in this review are published.

