## Supplemental material for "Efficacy and safety of TiNO-coated stents versus drug-eluting coronary stents. Systematic literature review and meta-analysis"

**Note: References used in this document are listed in the manuscript.**

**Abbreviations**

| | |
|---|---|
| ACS | Acute Coronary Syndrome |
| BAS | Bio-Active Stent |
| BMS | Bare-Metal Stent |
| DAPT | Dual Antiplatelet Therapy |
| CAD | Coronary Artery Disease |
| CD | Cardiac Death |
| CI | Confidence Intervals |
| DES | Drug-Eluting Stent |
| GRADE | Grading of Recommendations Assessment, Development and Evaluation |
| MACE | Major Adverse Cardiac Events |
| MI | Myocardial Infarction |
| N.S. / n.s. | Not Significant |
| N.A. | Not Applicable |
| N.R. | Not Reported |
| PCI | Percutaneous Coronary Interventions |
| PRISMA | Preferred Reporting Items for Systematic Reviews and Meta-Analyses |
| RCT | Randomized Clinical Trial |
| RR | Risk Ratio |
| SLR | Systematic Literature Review |
| ST | Stent Thrombosis |
| TD | Total Death meaning all-cause mortality |
| TiNOS | Titanium-Nitride Oxide coated-coronary Stent |
| TLR | Target Lesion Revascularization |
| TVR | Target Vessel Revascularization |
| OIS | Optimal Information Size |
| PICOS | Patient Intervention Comparator Outcome Study |

#### Part 1: Systematic review methods

##### Cochrane method for the analysis of study bias

Risk of bias in individual RCTs was assessed according to eight criteria proposed by the Cochrane Collaboration: (1) Random sequence generation (selection bias), (2) Allocation concealment (selection bias), (3) Blinding of outcome assessment (detection bias), (4) Incomplete outcome data (attrition bias), (5) Selective outcome reporting (reporting bias), (6) Performance bias was split into two components: (6a) blinding of operator versus (6b) blinding of participants and other personnel. The last item was (7) Other sources of bias.[16,30] The split in performance bias was based on the fact that operators could not be blinded while other personnel and other caregivers could.

##### GRADE rating of certainty of evidence

Each outcome was rated high, moderate, low or very low.[31-33] That rating combined the following five aspects: (1) Risk of bias, (2) inconsistency with respect to the degree of heterogeneity between studies, (3) indirectness between the evidence and the question to be addressed as defined according to PICOS, (4) publication bias and (5) imprecision related to evidence fragility considering the width of confidence intervals, Optimal Information Size "OIS" analysis related to the numbers of participants and cases, the threshold for benefit or harm and sensitivity analysis to redistribution of cases. OIS was defined as the amount of evidence that would have been required in a single RCT to test the hypotheses with the enough power. The amount of evidence expected in a meta-analysis should not be less than the OIS.[33,34]

The GRADE working group also uses a rule of thumb of a minimum of n = 300 to 500 events as an alternative if the OIS is not met. Both the OIS and this rule of thumb may be unattainable in rare events such as adverse events. Depending on the criticality of the safety events, the precision of the comparative risk of event may be rated as good if the 95% CI is narrow with boundaries far from 1.

**Heterogeneity estimation method**

**source: https://gdt.gradepro.org/app/handbook/handbook.html**

Heterogeneity is the variability in results. Inconsistency refers to an unexplained heterogeneity of results. True differences in the underlying treatment effect is likely when there are widely differing estimates of the treatment effect (i.e.) across studies. Explanations for heterogeneity, will be explored and if no plausible explanation is identified, the quality of evidence should be downgraded. Whether it is downgraded by one or two levels will depend on the magnitude of the inconsistency in the results.

Patient baseline the risk of adverse outcomes that the treatments are designed to prevent (e.g. death) presents variability so the risk differences and absolute risk reductions (ARR) in subpopulations tend to vary widely. Relative risk (RR) reductions, on the other hand, tend to be similar across subgroups, even if subgroups have substantial differences in baseline risk. Therefore, inconsistencies in effect size refer to relative measures (risk ratios and hazard ratios, which are preferred, or odds ratios).

Easily identifiable patient characteristics confidently permit classifying patients into subpopulations at appreciably different risk, absolute differences in outcome between intervention and control groups will differ substantially between these subpopulations. This may well warrant differences in recommendations across subpopulations, rather than downgrading the quality evidence for inconsistency in effect size.

There are a variety of criteria to assess heterogeneity when results cannot be pooled statistically as well as statistical methods to measure heterogeneity. Criteria to determine whether to downgrade for inconsistency can be applied when results are from more than one study and include:

- Wide variance of point estimates across studies (note: direction of effect is not a criterion for inconsistency)
- Minimal or no overlap of confidence intervals (CI), which suggests variation is more than what one would expect by chance alone.
- Cochran's Q-test of heterogeneity considered significant (reject the null hypothesis of no heterogeneity) if $p < 0.05$.
- The $I^2$ index *($I^2=(Q-df)/Q\times 100\%$, where $Q=\chi^2$ and df: corresponding degrees of freedom)* was calculated in order to overcome insufficient power of the Q-test.[35,36] The $I^2$ estimated the percentage of heterogeneity between study RRs that is not attributable to random error.[37,38] The rule of thumb proposed by GRADE: " < *40% may be low, 30-60% may be moderate, 50-90% may be substantial, 75-100% may be considerable".*

This protocol considered inconsistency to be

- not serious (low heterogeneity) if $I^2 < 50\%$ and Cochran' $Q \geq 0.05$ and if no confidence interval overlapped.

- Serious (moderate heterogeneity) if one only of those three criteria were met: if $I^2 \geq 50\%$ or Cochran' Q <0.05 or at least one confidence interval did not overlap with the others
- Very serious (substantial heterogeneity) if two or three of those criteria were met.

Low heterogeneity within a subgroup warranted a fixed-effect model, high heterogeneity a random effects model and moderate heterogeneity would require additional information to choose one or to rely mainly on the sensitivity analysis.

Risk of publication bias was assessed for each endpoint with a funnel plot and supported by Harbord's regression test calculated in STATA with the metabias command.[39-42]

**Assumptions and criteria used in this meta-analysis**

The assumption was made that a fixed-effect model was applicable within ACS and within the "other CAD" subgroups respectively, but not when the two subgroups were pooled. Publication bias was estimated for each endpoint separately with a funnel plot and Harbord's test. An asymmetrical funnel plot and a Harbord regression test for dichotomous endpoints with $p < 0.005$ indicated a significant risk of publication bias. Although both methods lack the power to detect publication bias when there are less than 10 studies, "no risk of publication bias detected" would be reported if such were the results of the two indicators as recommended in GRADE guidelines.

Generalizing the interpretation of a pooled outcome was attempted only if no significant publication bias relative to that outcome was detected or suspected.

Each pooled RR underwent a two-sided test of the null hypothesis (H0) that TiNOS and DES treatment arms presented the same risk of event *vs.* the alternative hypothesis (H1) that the risk of even was different. H0 was rejected in favor of H1 if the 95% CI of the RR did not include the no-effect value: 1.0.

The robustness of each outcome's pooled RR was assessed through sensitivity analysis, which consisted in iteratively recalculating the pooled RR after removing the input of one eligible RCT. As a result, the impact of each single RCT on the pooled RR of each outcome was quantified.

#### Part 2: Additional results of the meta-analysis

#### Table 1 Methods of the included RCTs

| Study | Population | Treatment | Endpoints & follow-up | Study design |
|---|---|---|---|---|
| TITAX-AMI<br>[NCT00495664]<br>Funding by institutions + Hexcath grant | Inclusion criteria: ACS: de novo lesions; NSTEMI & STEMI<br>Enrolment: 6 centers in Finland : Dec 2005 to Nov 2006 | DES: stainless steel stent platform, polyolefin polymer derivative, paclitaxel<br>TiNOS: TiNO coating, stainless steel platform | Primary: 12m MACE = cardiac death, recurrent myocardial infarction, or ischemia driven TLR<br>Stent thrombosis per ARC criteria<br>Planned: 1y through 5y. 12m & 5y completed | Non-inferiority in terms of 12m MACE. Two-sided significance test.<br>Sample size calculated to detect a minimum difference in true rates of d>=8%: α=5%, power =80%<br>Sample size: 400<br>1:1 randomization & assessor blinding |
| TIDE<br>[NCT00492908]<br>Funding by institution | Inclusion criteria: CAD with stenosis>=50% with stable or unstable angina pectoris or NSTEMI.<br>Enrolment: 3 centers: Switzerland June 2007 to Sept. 2008. | DES: phosphorylcholine coating, zotarolimus eluting, cobalt-chromium platform versus<br>TiNOS: TiNO coating, stainless steel platform | Primary: 6 to 8 month in-stent LLL.<br>Secondary: TD, CD, MI, clinically and non-clinically indicated TLR. TVR MACE & clinically indicated TLR MACE, ST per ARC criteria<br>Planned: 1y through 5y. 12m & 5y completed | Non-inferiority in terms of 6 to 8 month in-stent LLL. One-sided test.<br>H0: expected LLL DES= 0.61±0.46mm vs. TiNOS=0.55±0.63mm<br>non-inferiority margin d=0.15mm<br>α=5%, power>=80%<br>Sample size: 234<br>1 randomization & assessor blinding |
| TITANIC-XV<br>[NCT01510509]<br>Funding: not disclosed | Inclusion criteria: de novo coronary lesion with stenosis>=50% in diabetic patients.<br>Enrolment period: 8 centers in Spain & Finland Jan. 2009 to Oct. 2011. | PCI with<br>DES: polyvinylidene fluoride-co-hexafluoropropylene coating, everolimus eluting, cobalt-chromium platform versus<br>TiNOS: TiNO coating, stainless steel platform | Primary angiographic: 6 to 8 month in-stent LLL.<br>Primary clinical: at 12 months of FU: MACE-1 TD, non-fatal MI, stroke, or clinically indicated TVR. **used as proxy for TLR MACE in this in the meta-analysis.**<br>Secondary: at 12-m FU: TD, TLR, TVR, repeat revascularization of non- target vessel, composite end point of death, nonfatal AMI, stroke, or repeat revascularization of any site (composite = MACE-2); ST per ARC; and clinical restenosis.<br>Planned: 1y through 2y. 12m & 2y completed but 1y published only | Single sided superiority test in terms of MACE 1.<br>H0: No difference in MACE 1 between DES and TiNOS<br>HA: 23% MACE 1 with TiNOS and 8% with DES, i.e. absolute risk reduction ARR=15%<br>α=5%, power=80%<br>Randomization: 1 DES : 1 TiNOS<br>Assessor blinding. |
| BASE-ACS<br>[NCT00819923]<br>Funding by institutions + Hexcath grant | Inclusion criteria: ACS with >= 1 de novo stenosis>50% (visual estimation) in a native coronary artery or bypass graft in the presence of NSTEMI or STEMI or unstable angina.<br>Enrolment: 14 centers in Finland, Belgium, UK, Spain, Switzerland, Indonesia: Nov 2008 to end N.R. | DES: polyvinylidene fluoride-co-hexafluoropropylene coating, everolimus eluting, cobalt-chromium platform versus<br>TiNOS: TiNO coating, stainless steel platform | Primary: 12m MACE = cardiac death or non-fatal myocardial infarction, or ischemia driven TLR<br>Secondary: all-cause death, composite of cardiac death or non-fatal MI and definite ST per ARC at 12-month<br>Planned: 1y through 5y. 12m & 5y completed | Non-inferiority in terms of 12m MACE:<br>One-sided tests.<br>H0: expected MACE rate DES=9.2%, TiNOS > 9.2% + non-inferiority margin d=5%<br>HA: MACE rate difference TiNOS – DES< d<br>α=5%, power>=90%, sample size: 800<br>Randomization: 1 DES : 1 TiNOS |
| TIDES-ACS<br>[NCT02049229]<br>Funding by institutions + Hexcath grant | Inclusion criteria: ACS: de novo coronary lesion with NSTEMI & STEMI<br>Exclusion criteria: life<br>Enrolment period: Jan 2014 to Aug 2016<br>12 centers: Finland, Netherland, Spain, Belgium, Luxemburg, Switzerland | DES: PLGA absorbable coating, everolimus eluting, platinum-chromium platform versus<br>TiNOS: TiNO coating, cobalt-chromium platform | Primary: 12m MACE = CD or non-fatal MI, or ischemia driven TLR<br>Co-primary endpoint: 18-month = composite of CD, any MI, major bleeding per BARC<br>ST per ARC<br>Secondary: at 12m: composite of CD, MI, ST, TLR; cardiac death or MI; cardiac death; MI; ST; all-cause death; TLR; TVR; major bleeding<br>Planned: 1y through 5y. 12m & 18m completed & 5y on-going | Non-inferiority in terms of 12m MACE:<br>After interim analysis:<br>H0: expected MACE rate DES=7%, TiNOS>7% + non-inferiority margin d=3.5%<br>HA: MACE rate difference TiNOS – DES< d<br>α=5%, power=80%<br>Sample size: 1476<br>Randomization: 1 DES : 2 TiNOS |

### Table 2 Number of patients with events – ITT

| | OUTCOME STUDY: | TITAX-AMI | TIDE | TITANIC-XV | BASE-ACS | TIDES-ACS |
|---|---|---|---|---|---|---|
| Stent | ACS subgroup data | Yes | No | No | Yes | Yes |
| | Enrolled | 214 | 152 | 83 | 417 | 989 |
| | Modified device-oriented MACE 1-y | 22 | 27 | (7+1 [b]) 8 | 40 | 62 |
| | Modified device-oriented MACE 5-y | 35 | 33 | N.A. | 57 | Expected |
| | CD or MI 1-y | 9 | 8 | 1 [f] | 16 | 23 |
| | CD or MI 5-y | 18 | 12 | N.A. | 30 | Expected |
| | Non-fatal MI 1-y | (9-1 [e]) 8 | (8–0 [e]) 8 | 1 | 9 | (23-5 [e]) 18 |
| | Non-fatal MI 5-y | (18-4 [e]) 16 | (12–5 [e]) 7 | N.A. | 21 | Expected |
| | CD 1-y | 1 | 0 | 0 | 8 | 5 |
| | CD 5-y | 4 | 5 | N.A. | 11 | Expected |
| | TLR 1-y (clinically driven) | 18 [d] | 22 | N.A. (assumed 7) | 27 | 53 |
| | TLR 5-y (clinically driven) | 24 | 24 | N.A. | 33 | Expected |
| | probable or definite ST 1-y | 1 | 1 | 0 | 5 | 11 |
| | probable or definite ST 5-y | 2 [a] | 1 | N.A. | (5+1 [a]) 6 | Expected |
| TiNOS | definite ST 1-y | 1 | 1 | 0 | 3 | 10 |
| | definite ST 5-y | 2 | 1 | N.A. | 4 | Expected |
| | Recurrent MI 1-y | 9 | 8 | N.A. | N.A. | 18 |
| | Recurrent MI 5-y | 18 | 8 | N.A. | N.A. | Expected |
| | TD 1-y | 5 | 1 | 0 | 15 | 9 |
| | TD 5-y | 19 | 13 | N.A | 31 | Expected |
| | TLR 1-y (any) | 20 | 26 | 7 | N.A. | N.A. |
| | TLR 5-y (any) | N.A. | N.A. | N.A. | N.A. | Expected |
| | TVR 1-y (clinically driven) | N.A. | 27 | N.A. | N.A. | 65 |
| | TVR 5-y (clinically driven) | N.A. | 33 | N.A. | N.A. | Expected |
| | TVR 1-y (any) | N.A. | 31 | 11 | N.A. | N.A. |
| | TVR 5-y (any) | N.A. | N.A. | N.A. | N.A. | Expected |
| | Lost to follow-up 1-y | 0 | 0 | N.A. | 3 | 7 |
| | Lost to follow-up 5-y | 3 | N.A. | N.A. | 29 [c] | Expected |
| | Enrolled | 211 | 150 | 90 | 410 | 502 |
| | Modified device-oriented MACE 1-y | 27 | 21 | (3+2 [b]) 5 | 37 | 35 |
| | Modified device-oriented MACE 5-y | 53 | 28 | N.A. | 69 | Expected |
| | CD or MI 1-y | 18 | 10 | 2 [f] | 26 | 25 |
| | CD or MI 5-y | 42 | 14 | N.A. | 48 | Expected |
| | Non-fatal MI 1-y | (18-4 [e] ) 14 | (10-0) 10 | 2 | 24 | (25-8 [e] ) 17 |
| | Non-fatal MI 5-y | (42-12 [e] ) 30 | (14–4 [e]) 10 | N.A. | 37 | Expected |
| | CD 1-y | 4 | 0 | 0 | 4 | 8 |
| | CD 5-y | 12 | 4 | N.A. | 13 | Expected |
| DES | TLR 1-y (clinically driven) | 9 [d] | 13 | N.A. (assumed 3) | 20 | 17 |
| | TLR 5-y (clinically driven) | 23 | 19 | N.A. | 37 | Expected |
| | probable or definite ST 1-y | 8 | 0 | 0 | 11 | 14 |
| | probable or definite ST 5-y | 15 [a] | 0 | N.A. | (1+5 [a]) 16 | Expected |
| | definite ST 1-y | 1 | 0 | 0 | 9 | 10 |
| | definite ST 5-y | 15 | 0 | N.A. | 14 | Expected |
| | Recurrent MI 1-y | 17 | 10 | N.A. | N.A. | 23 |
| | Recurrent MI 5-y | 38 | 11 | N.A. | N.A. | Expected |
| | TD 1-y | 6 | 1 | 0 | 10 | 13 |
| | TD 5-y | 22 | 8 | N.A | 30 | Expected |
| | TLR 1-y (any) | 15 | 17 | 3 | N.A. | N.A. |

| | | | | | | |
|---|---|---|---|---|---|---|
| | TLR 5-y (any) | N.A. | N.A. | N.A. | N.A. | Expected |
| | TVR 1-y (clinically driven) | N.A. | 20 | N.A. | N.A. | 21 |
| | TVR 5-y (clinically driven) | N.A. | 31 | N.A. | N.A. | Expected |
| | TVR 1-y (any) | N.A. | 23 | 3 | N.A. | N.A. |
| | TVR 5-y (any) | N.A. | N.A. | N.A. | N.A. | Expected |
| | Lost to follow-up 1-y | 0 | 2 | N.A. | 3 | 4 |
| | Lost to follow-up 5-y | 7 | N.A. | N.A. | 28[c] | Expected |

Notes:

[a] 5-year Probable or definite estimated using 1-year Probable or definite + new cases of definite at 5-year follow-up

[b] MACE estimated as TLR + non-fatal MI assuming no overlap, given death = 0

[c] Communicated by primary investigator

[d] Due to in-stent restenosis

[e] Non-fatal MI = Cardiac death OR MI – Cardiac death

[f] Cardiac death or MI in TITANIC-XV: Cardiac death = 0 and number of non-fatal MI used as best estimate of number of MIs.

- N.A. Not applicable

- TITANIC-XV reports patient-oriented MACE only using TVR instead of TLR.

-BASE-ACS stent thrombosis per ARC definition: probable or definite at 1-year follow-up, and definite-only at 5-year follow-up.

- Recurrent MI is described as non-fatal in BASE-ACS, TIDES-ACS, and TITANIC-XV. It is not specified as fatal or non-fatal in TITAX-AMI and TIDE. The reviewers assume that patients with fatal MI are relatively few compared to patients with non-fatal MI. This causes minimum indirectness in the pooled analysis. Moreover, the RR of CD or MI, which includes all MIs is consistent with the RR of recurrent MI.

#### PUBLICATION BIAS ANALYSIS

**Figure 1a Funnel plot - MACE RR 1-year– All CAD**

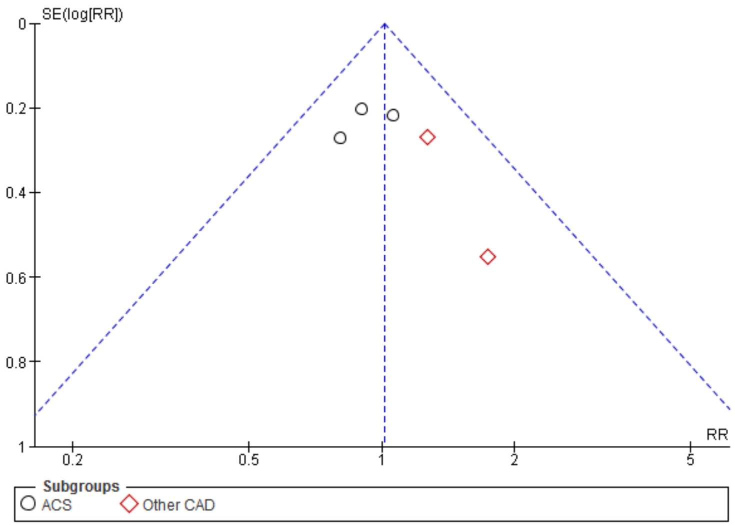


Non-significant Harbord test: p = 0.263

**Figure 1b Funnel plot RR – CD or MI 1-year– All CAD**

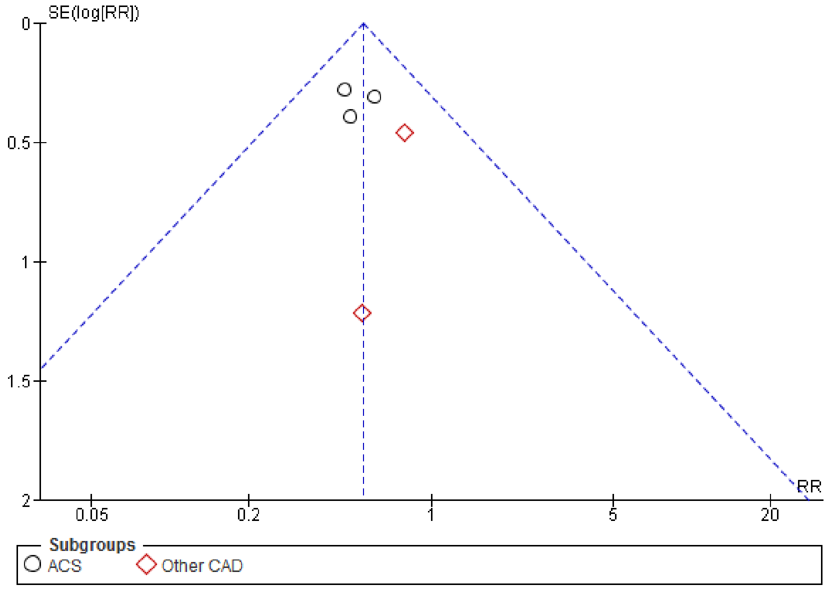


Non-significant Harbord test: p = 0.660

**Figure 1c Funnel plot RR – non-fatal MI 1-year– All CAD**

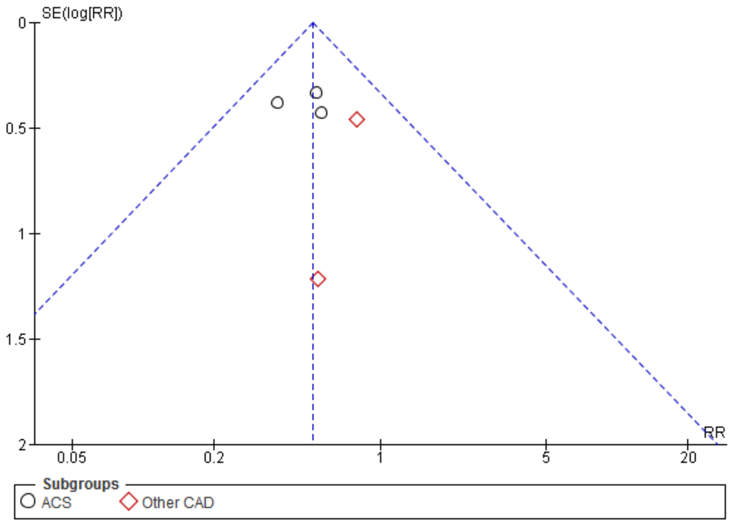


non-significant Harbord test: p = 0.678

**Figure 1d Funnel plot RR – CD 1-year– All CAD**

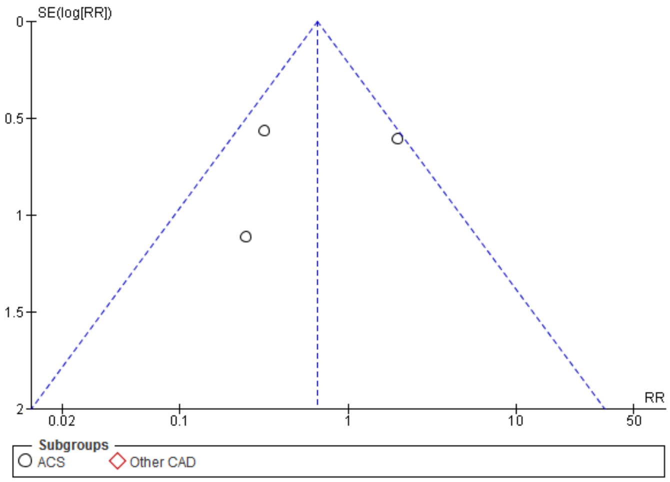


Non-significant Harbord test: p = 0.727

**Figure 1e Funnel plot RR – clinically driven TLR 1-year– All CAD**

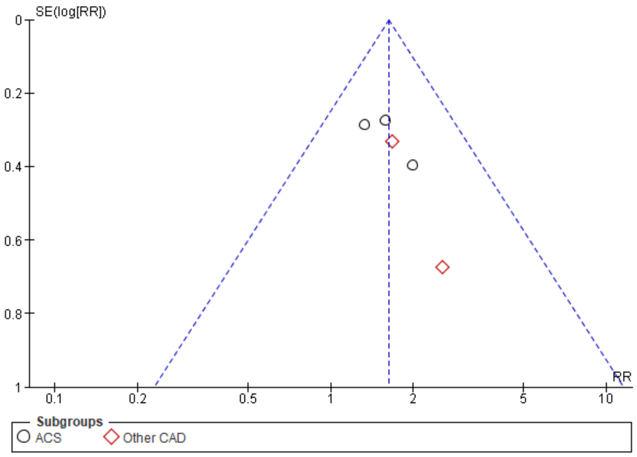


Non-significant Harbord test: p = 0.086

**Figure 1f Funnel plot RR – probable or definite ST 1-year– All CAD**

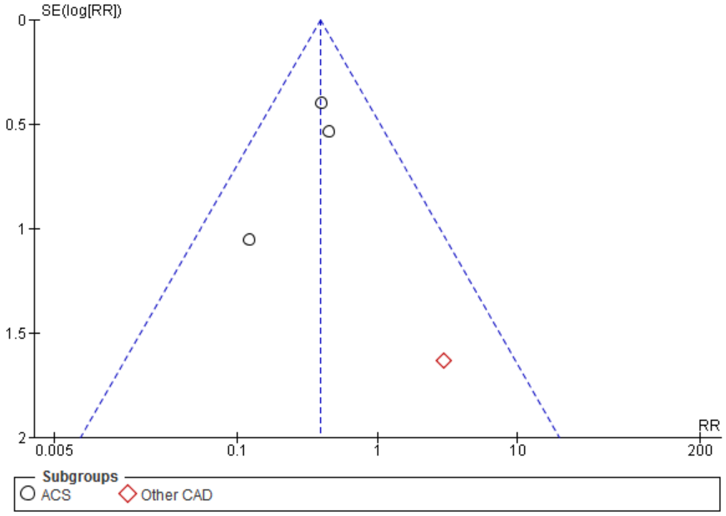


Non-significant Harbord test: p = 0.373

**Figure 1g Funnel plot RR – TD 1-year– All CAD**

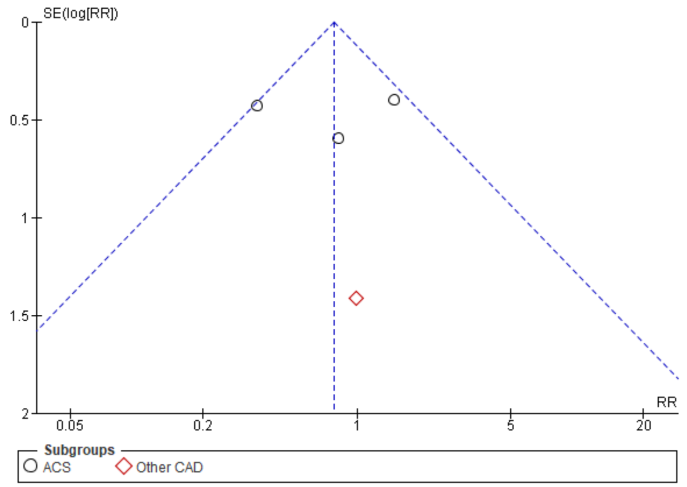


non-significant Harbord test: p = 0.962

**Figure 2a Risk of bias of individual studies**

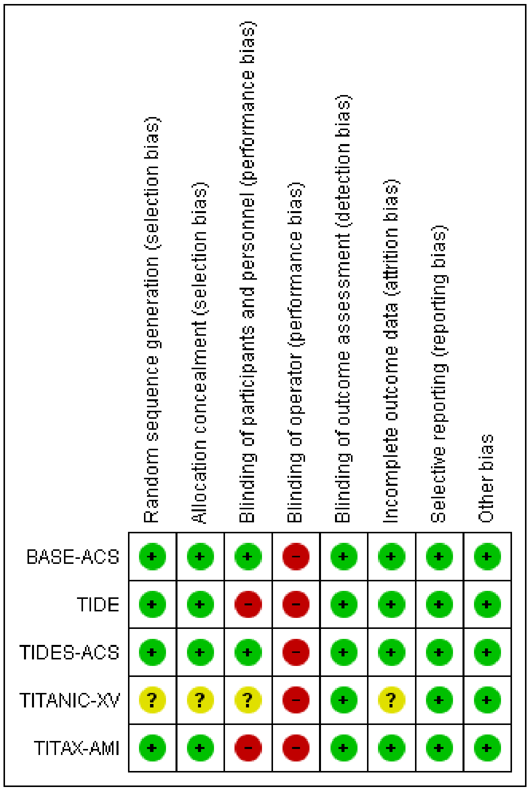


**Figure 2b Risk of bias across the studies**

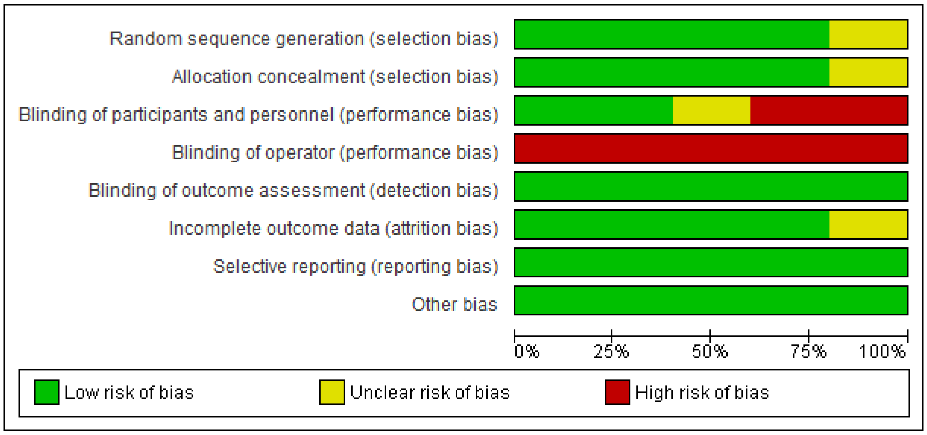

#### Figure 3a Forest plot - RR – CD or MI 1-year

| MI or CD 1-year<br>Study or Subgroup | TiNOS<br>Events | Total | DES<br>Events | Total | Weight | Risk Ratio<br>M-H, Fixed, 95% CI | Year |
|---|---|---|---|---|---|---|---|
| **ACS** | | | | | | | |
| TITAX-AMI | 9 | 214 | 18 | 211 | 20.3% | 0.49 [0.23, 1.07] | 2008 |
| BASE-ACS | 16 | 417 | 26 | 410 | 29.3% | 0.61 [0.33, 1.11] | 2012 |
| TIDES-ACS | 23 | 989 | 25 | 502 | 37.1% | 0.47 [0.27, 0.81] | 2020 |
| **Subtotal (95% CI)** | | **1620** | | **1123** | **86.6%** | **0.52 [0.36, 0.75]** | |
| Total events | 48 | | 69 | | | | |
| Heterogeneity: Chi² = 0.40, df = 2 (P = 0.82); I² = 0% | | | | | | | |
| Test for overall effect: Z = 3.54 (P = 0.0004) | | | | | | | |
| **Other CAD** | | | | | | | |
| TIDE | 8 | 152 | 10 | 150 | 11.2% | 0.79 [0.32, 1.95] | 2011 |
| TITANIC-XV | 1 | 83 | 2 | 90 | 2.1% | 0.54 [0.05, 5.87] | 2011 |
| **Subtotal (95% CI)** | | **235** | | **240** | **13.4%** | **0.75 [0.32, 1.74]** | |
| Total events | 9 | | 12 | | | | |
| Heterogeneity: Chi² = 0.08, df = 2 (P = 0.77); I² = 0% | | | | | | | |
| Test for overall effect: Z = 0.67 (P = 0.50) | | | | | | | |
| **Total (95% CI)** | | **1855** | | **1363** | **100.00%** | **0.55 [0.39, 0.77]** | |
| Total events | 57 | | 81 | | | | |
| Heterogeneity: Chi² = 1.12, df = 4 (P = 0.89); I² = 0% | | | | | | | |
| Test for overall effect: Z = 3.52 (P = 0.0004) | | | | | | | |
| Test for subgroup differences: Chi² = 0.61, df = 1 (P = 0.43), I² = 0% | | | | | | | |

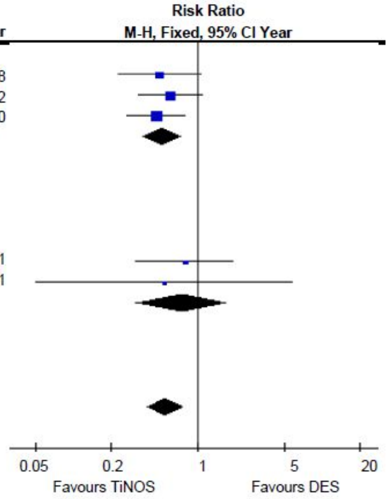


#### Figure 3b Forest plot - RR - TD 1-year

| TD 1-year<br>Study or Subgroup | TiNOS<br>Events | Total | DES<br>Events | Total | Weight | Risk Ratio<br>M-H, Fixed, 95% CI | Year |
|---|---|---|---|---|---|---|---|
| **ACS** | | | | | | | |
| TITAX-AMI | 5 | 214 | 6 | 211 | 17.6% | 0.82 [0.25, 2.65] | 2008 |
| BASE-ACS | 15 | 417 | 10 | 410 | 29.3% | 1.47 [0.67, 3.24] | 2012 |
| TIDES-ACS | 9 | 989 | 13 | 502 | 50.2% | 0.35 [0.15, 0.82] | 2020 |
| **Subtotal (95% CI)** | | **1620** | | **1123** | **97.1%** | **0.78 [0.47, 1.27]** | |
| Total events | 29 | | 29 | | | | |
| Heterogeneity: Chi² = 5.95, df = 2 (P = 0.05); I² = 66% | | | | | | | |
| Test for overall effect: Z = 1.01 (P = 0.31) | | | | | | | |
| **Other CAD** | | | | | | | |
| TIDE | 1 | 152 | 1 | 150 | 2.9% | 0.99 [0.06, 15.63] | 2011 |
| TITANIC-XV | 0 | 83 | 0 | 90 | | Not estimable | 2011 |
| **Subtotal (95% CI)** | | **235** | | **240** | **2.9%** | **0.99 [0.06, 15.63]** | |
| Total events | 1 | | 1 | | | | |
| Heterogeneity: Not applicable | | | | | | | |
| Test for overall effect: Z = 0.01 (P = 0.99) | | | | | | | |
| **Total (95% CI)** | | **1855** | | **1363** | **100.00%** | **0.66 [0.33, 1.31]** | |
| Total events | 30 | | 30 | | | | |
| Heterogeneity: Chi² = 5.98, df = 3 (P = 0.11); I² = 50% | | | | | | | |
| Test for overall effect: Z = 1.00 (P = 0.32) | | | | | | | |
| Test for subgroup differences: Chi² = 0.03, df = 1 (P = 0.87), I² = 0% | | | | | | | |

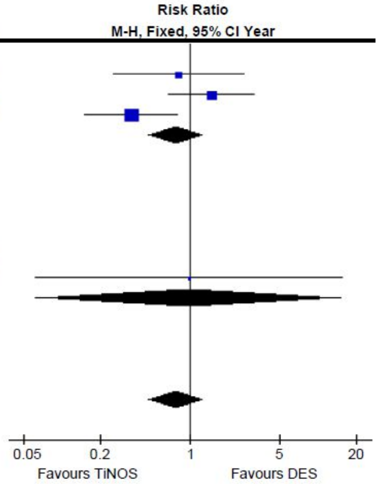

#### GRADE assessment

#### Table 3a GRADE certainty of evidence assessment - 1-year follow-up in ACS

| Outcome | Risk of bias | Inconsistency | Indirectness | Imprecision | Publication bias | Overall certainty of evidence |
|---|---|---|---|---|---|---|
| Device oriented MACE | not serious [a] | not serious [b] | not serious [c] | not serious [d] | none | ⊕⊕⊕⊕ HIGH |
| CD or MI | not serious [a] | not serious [b] | not serious [e] | serious [f] | none | ⊕⊕⊕◯ MODERATE |
| Clinically driven TLR | not serious [a] | not serious [b] | not serious [g] | very serious [h] | none | ⊕⊕◯◯ LOW |
| Non-fatal MI | not serious [a] | not serious [b] | not serious [i] | serious [j] | none | ⊕⊕⊕◯ MODERATE |
| CD | not serious [a] | very serious [k] | not serious [l] | very serious [m] | none | ⊕◯◯◯ VERY LOW |
| Probable or definite ST | not serious [a] | not serious [b] | not serious [n] | very serious [o] | none | ⊕⊕◯◯ LOW |
| TD | not serious [a] | serious [k,p] | not serious [q] | very serious [r] | none | ⊕◯◯◯ VERY LOW |

**Explanations**

a. The risk of bias common to all RCTs was operator knowledge of the type of stent. However, given the absence of significant differences in baseline and procedural data, that risk seems to have had no effect. Other potential risks of bias were occasional in some RCTs but the sensitivity analysis shows that bias related to individual RCTs has little influence on pooled outcomes. The study with the highest risk of bias has the smallest relative weight and does not report outcomes in ACS or at 5-year follow-up, so the impact the potential risk of that trial is limited.

b. Heterogeneity within that subgroup was low given the overlap of all confidence intervals, Cochran Q-test $p>0.05$ and $I^2< 40\%$.

c. The 3 ACS RCTs report modified device-oriented MACE. Assuming half of non-fatal MIs were in nontarget vessels (i.e. 45 cases), the proportionality would lead to 27 fewer cases with TiNOS and 18 with DES. If these numbers were removed from the count of MACE, the RR would be 0.98 [0.73, 1.30], which does not change the non-inferiority conclusion.

d. The 95% CI of the RR of MACE included 1.0 with a lower bound slightly < 0.75 favorable to TiNOS and a length slightly > 0.5. This result is robust to sensitivity analysis. A hypothetical single RCT to demonstrate non-inferiority of TiNOS compared to DES, 1:1 randomization, 2-sided test, $\alpha = 2.5\%$, power = 90%, reference rate of events in the 3 trial DES arms 99/1123 = 0.088 assuming a non-inferiority margin delta = 0.035 based on a targeted 30% RRR requires a total sample size of n = 2,760 patients, which is 1.01 times the pooled sample size. Considering the total number of events observed is 124+99 = 223 reaching the GRADE rule of thumb of 300 events overall requires n = 2760 * 300/223 = 3,713 patients to meet the OIS criterion. However, with the current sample size, a minimum of 20 events (9% of observed events) should be redistributed from the DES arm to the TiNOS arm for the CI to reach non-significance (RR 1.42 [1.06, 1.91]).

e. Publications usually do not indicate if the MI is related to the target vessel or not, but this review assumes that recurrent MIs related to non-target vessels at 1-year follow-up should be in equal proportion in the two treatment arms.

f. The 95% CI of the RR of ST is significantly < 1.0 in favour of TiNOS with an upper-bound = 0.75 and a length < 0.5. This result is robust to sensitivity analysis. A hypothetical single RCT, 1:1 randomization, 2-sided test, α = 5%, power = 80%, reference rate of events in the 3 trial DES arms 69/1123 = 0.061 assuming delta = 0.018 based on a targeted 30% RRR requires a total sample size of n = 4,788 patients, which is 1.75 times the pooled sample size. Considering the total number of events observed is 48+69 = 117 reaching the GRADE rule of thumb of 300 events overall requires n = 4788 * 300/117 = 12,277 patients to meet the OIS criterion. With the current sample size, a minimum of 9 event (7.7% of observed events) should be redistributed from the DES arm to the TiNOS arm for the CI to reach non-significance (RR 0.71 [0.49, 1.02]). This result is imprecise and fragile.

g. TLR defined as clinically-driven confirmed in all three ACS trials

h. The 95% CI of the RR of TLR is significantly > 1.0 in favour of DES with a lower bound < 1.25 and a length > 1. This result is not robust to sensitivity analysis. A hypothetical single RCT, 1:1 randomization, 2-sided test, α = 5%, power = 80%, reference rate of events in the 3 trial DES arms 46/1123 = 0.041 assuming delta = 0.012 based on a targeted 30% RRR requires a total sample size of n = 7,308 patients, which is 2.66 times the pooled sample size. Considering the total number of events observed is 98+46 = 144 reaching the GRADE rule of thumb of 300 events overall requires n = 7308 * 300/144 = 15,225 patients to meet the OIS criterion. With the current sample size, a minimum of 3 events (2.1% of observed events) should be redistributed from the TiNOS arm to the DES arm for the CI to reach non-significance (RR 1.40 [1.00, 1.97]). This result is imprecise and fragile.

i. Recurrent MI is described as non-fatal in the three RCTs and PK confirms that those included non-fatal MIs in nontarget vessels. One can reasonably assume the index stent does not affect those events. Ninety non-fatal MIs are reported. Assuming half of them are related to nontarget vessels (i.e., 45 cases), proportionality with sample size would lead to 27 fewer cases with TiNOS and 18 with DES, which would result in an RR of 0.19 [0.09, 0.42]. The inclusion of nontarget MIs thus results in a dilution that is favorable to DES.

j. The 95% CI of the RR of non-fatal MI is significantly < 1.0 in favour of TiNOS with an upper-bound < 0.75 and a length < 0.5. This result is robust to sensitivity analysis. A hypothetical single RCT, 1:1 randomization, 2-sided test, α = 5%, power = 80%, reference rate of events in the 3 trial DES arms 55/1123 = 0.049 assuming delta = 0.015 based on a targeted 30% RRR requires a total sample size of n = 6,070 patients, which is 2.21 times the pooled sample size. Considering the total number of events observed is 35+55 = 90 reaching the GRADE rule of thumb of 300 events overall requires n = 6070 * 300/90 = 20,233 patients to meet the OIS criterion. With the current sample size, a minimum of 8 events (8.9% of observed events) should be redistributed from the DES arm to the TiNOS arm for the CI to reach non-significance (RR 0.68 [0.44, 1.05]). This result is imprecise and fragile.

k. Heterogeneity within that subgroup was high given the absence of overlap of confidence intervals, the significant Cochran Q-test p=0.005 and $I^2 \geq 80\%$.

l. The definition of CD was assumed to be similar across the 3 ACS RCTs. This uniform reporting method across the three trials was confirmed by PK.

m. The 95% CI of the RR of CD is non-significant with a length > 0.5. Sensitivity analysis supports no association. A hypothetical single RCT, 1:1 randomization, 2-sided test, α = 5%, power = 80%, reference rate of events in the 3 trial DES arms 16/1123 = 0.014 assuming delta = 0.004 based on a targeted 30% RRR requires a total sample size of n = 21,484 patients, which is 7.83 times the pooled sample size. Considering the total number of events observed is 14+16 = 30 reaching the GRADE rule of thumb of 300 events overall requires n = 21484 * 300/30 = 214,840 patients to meet the OIS criterion. With the current sample size, a minimum of 3 events (10% of observed events) should be redistributed from the TiNOS arm to the DES arm for the CI to reach significance (RR 0.45 [0.23 0.90]). This result is imprecise and fragile.

n. ARC criteria used in all studies. Probable or definite stent thrombosis at 1-y FU.

o. The 95% CI of the RR of ST is significantly < 1.0 in favour of TiNOS with an upper-bound > 0.75 and a length > 0.5. This result is robust to sensitivity analysis. A hypothetical single RCT, 1:1 randomization, 2-sided test, $\alpha = 5\%$, power = 80%, reference rate of events in the 3 trial DES arms 20/1123 = 0.018 assuming delta = 0.005 based on a targeted 30% RRR requires a total sample size of n = 17136 patients, which is 6.25 times the pooled sample size. Considering the total number of events observed is 14+20 = 34 reaching the GRADE rule of thumb of 300 events overall requires n = 17136 * 300/34 = 151,200 patients to meet the OIS criterion. With the current sample size, a minimum of 1 event (2.9% of observed events) should be redistributed from the DES arm to the TiNOS arm for the CI to reach non-significance (RR 0.52 [0.26, 1.03]). This result is imprecise and fragile.

p. Heterogeneity within that subgroup was moderate given the overlap of the confidence intervals, but the significant Cochran Q-test p=0.05 and $I^2 \geq 60\%$.

q. The definition of TD was assumed to be similar across the RCTs so that the risk indirectness was rated not serious. The fact that the sensitivity analysis showed that the removal of BASE-ACS from the pooled analysis led to a significant confidence interval in favor of TiNOS did not appear to be related to differences in TD definition.

r. The 95% CI of the RR of TD is non-significant with a length > 0.5. Sensitivity analysis supports no association. A hypothetical single RCT, 1:1 randomization, 2-sided test, $\alpha = 5\%$, power = 80%, reference rate of events in the 3 trial DES arms 29/1123 = 0.026 assuming delta = 0.008 based on a targeted 30% RRR requires a total sample size of n = 11,740 patients, which is 4.28 times the pooled sample size. Considering the total number of events observed is 29+29 = 58 reaching the GRADE rule of thumb of 300 events overall requires n = 11740 * 300/58 = 60,724 patients to meet the OIS criterion. With the current sample size, a minimum of 4 events (6.9% of observed events) should be redistributed from the TiNOS arm to the DES arm for the CI to reach significance (RR 0.60 [0.37 0.97]). This result is imprecise and fragile.

#### Table 3b GRADE Summary of findings - TiNOS vs. DES in ACS at 1-year follow-up

| Outcome | Anticipated absolute effects (95% CI) | | | Certainty of evidence | Interpretation |
|---|---|---|---|---|---|
| | DES | TINOS | Difference (TiNOS – DES) | | |
| device-oriented MACE | 8.8% | 8.2% [6.3 to 10.6] | 0.6% fewer [2.5 fewer to 1.8 more] | HIGH [a,b,c,d] | The incidence of device-oriented MACE is not associated with the type of stent. The level of certainty of the underlying evidence is high. |
| CD or MI | 6.1% | 3.2% [2.2 to 4.6] | 2.9% fewer [3.9 to 1.5 fewer] | MODERATE [a,b,e,f] | The incidence of CD or MI is likely to be lower with TiNOS than with DES. The level of certainty of the underlying evidence is moderate. |
| clinically driven TLR | 4.1% | 6.3% [4.5 to 9] | 2.3% more [0.4 to 4.9 more] | LOW [a,b,g,h] | The incidence of TLR is likely to be higher with TiNOS than with DES. The level of certainty is low. |
| non-fatal MI | 4.9% | 2.4% [1.5 to 3.6] | 2.5% fewer [3.4 to 1.3 fewer] | MODERATE [a,b,i,j] | The incidence of non-fatal MI is likely to be lower with TiNOS than with DES. The level of certainty of the underlying evidence is moderate. |
| CD | 1.4% | 0.9% [0.5 to 1.9] | 0.5% fewer [1 to 0.4 more] | VERY LOW [a,k,l,m] | No association between the incidence of CD and the type of stent is detected. The level of certainty of the underlying evidence is very low. |
| probable or definite ST | 2.9% | 1.0% [0.6 to 1.9] | 1.9% fewer [2.4 to 1.1 fewer] | LOW [a,b,n,o] | The incidence of probable or definite ST is likely to be lower with TiNOS than with DES. The level of certainty of the underlying evidence is low. |
| TD | 2.6% | 2.0% [1.2 to 3.3] | 0.6% fewer [1.4 fewer to 0.7 more] | VERY LOW a,k,p,q,r | No association between the incidence of TD and the type of stent is detected. The level of certainty of the underlying evidence is very low. |

- CI: Confidence interval; RR: Risk ratio
- The risk with TiNOS and its 95% CI is based on the assumed risk with DES and the RR and its 95% CI.

GRADE Working Group grades of evidence

- High certainty: Very confident that the true effect lies close to that of the estimate.
- Moderate certainty: Moderately confident in the effect estimate: The true effect is likely to be close to the estimate of the effect, but there is a possibility that it is substantially different
- Low certainty: Confidence in the effect estimate is limited: The true effect may be substantially different from the estimate.
- Very low certainty: Very little confidence in the effect estimate: The true effect is likely to be substantially different from the estimate.
