## Supplementary material for "Efficacy and safety of TiNO-coated stents versus drug-eluting coronary stents. Systematic literature review and meta-analysis": PRISMA checklist

### PRISMA 2009 Checklist

Template from: Moher D, Liberati A, Tetzlaff J, Altman DG, The PRISMA Group (2009). Preferred Reporting Items for Systematic Reviews and Meta-Analyses: The PRISMA Statement. PLoS Med 6(7): e1000097. doi:10.1371/journal.pmed1000097

| Section/topic | # | Checklist item (location in report: Last line of each section, bold.) |
|---|---|---|
| **TITLE** | | |
| Title | 1 | *Identify the report as a systematic review, meta-analysis, or both.*<br>Efficacy and safety of percutaneous coronary interventions using titanium-nitride-oxide coated bioactive stents versus drug-eluting stents in coronary artery disease. A systematic review and meta-analysis.<br>**Article: cover page.** |
| **ABSTRACT** | | |
| Structured summary | 2 | *Provide a structured summary including, as applicable: background; objectives; data sources; study eligibility criteria, participants, and interventions; study appraisal and synthesis methods; results; limitations; conclusions and implications of key findings; systematic review registration number.*<br>- Objectives: To compare the efficacy and safety of PCI with TiNOS versus DES at 1 and 5-year follow-up, in all CAD including acute coronary syndrome (ACS).<br>- Methods: A prospective systematic literature review and meta-analysis of RCTs were conducted according to published methods (Cochrane, PRISMA, GRADE). Medline, Embase, Cochrane database, Web of Science were searched on March 08, 2018. Fixed-effects Mantel-Haenszel pooled risk ratios (RR), TiNOS over DES, with 95% confidence intervals (CI) were computed with sensitivity analysis.<br>- Results: Five RCTs were eligible with n = 1,855 TiNOS vs. n = 1,363 DES at 1-year and n = 783 vs. n = 771 at 5-year follow-up. One-year RRs in ACS patients: Major Adverse Cardiac Events (MACE) 0.93 [0.72, 1.20], recurrent myocardial infarction (MI) 0.42 [0.28, 0.63], probable or definite stent thrombosis (ST): 0.35 [0.20, 0.64]. Estimates were robust to sensitivity analysis. Certainty of evidence was high in MACE, moderate in MI and ST due to the limited number of cases. Data were insufficient to draw conclusions about stable CAD patients and 5-year outcomes.<br>- Conclusions: This meta-analysis showed in ACS patients, similar risk of MACE with TiNOS and DES, at 1-year follow-up, and suggested a lower risk of MI and ST with TiNOS than with DES. These two safety signals in MI and ST with the current use of DES require further investigations.<br>**Article: Abstract** |
| **INTRODUCTION** | | |
| Rationale | 3 | *Describe the rationale for the review in the context of what is already known.*<br>Percutaneous coronary interventions (PCI) with drug-eluting stents (DES) is the standard of care in Coronary Artery Disease (CAD), including Acute Coronary Syndrome (ACS). The use of DES carries a risk complications.<br>Titanium-nitride-oxide coated coronary stents (TiNOS), also designated “bioactive stents” (BAS) have a pharmacologically inactive, non-absorbable coating. Preclinical data has shown less neointimal hyperplasia with TiNOS than with bare-metal stents (BMS). Several trials comparing TiNOS with DES have been conducted but no systematic review of that evidence has been published so far.<br>**Article: Objectives** |

| | | |
|---|---|---|
| Objectives | 4 | *Provide an explicit statement of questions being addressed with reference to participants, interventions, comparisons, outcomes, and study design (PICOS).*<br>Question specification per PICOS:<br>- Patients: CAD including ACS and other forms of CAD such as stable angina pectoris.<br>- Intervention: PCI using TiNOS and the Comparator was PCI using DES.<br>- Outcomes: Device-oriented Major Adverse Cardiac Events (device-oriented MACE), recurrent myocardial infarction (MI), cardiac death (CD), clinically driven target lesion revascularization (TLR), stent thrombosis (ST), and all-cause mortality ("total death – TD") at 1-year and 5-year follow-up.<br>- Study methods were randomized controlled clinical trials (RCTs).<br>**Article: Participants – Interventions - Outcome measures.** |
| **METHODS** | | |
| Protocol and registration | 5 | *Indicate if a review protocol exists, if and where it can be accessed (e.g., Web address), and, if available, provide registration information including registration number.*<br>Protocol registered in PROSPERO N° CRD42018090622<br>Access: https://www.crd.york.ac.uk/prospero/display_record.php?RecordID=90622<br>**Article: Methods.** |
| Eligibility criteria | 6 | *Specify study characteristics (e.g., PICOS, length of follow-up) and report characteristics (e.g., years considered, language, publication status) used as criteria for eligibility, giving rationale.*<br>Study inclusion criteria: First-hand clinical evidence with prospective inclusion; patients with CAD treated with coronary PCI; implantation of either TiNOS or DES after random allocation; target outcomes reported at 1-year and/or 5-year follow-up; outcomes reported as the number of patients with an event or their rate along with the corresponding sample size.<br>Study exclusion criteria: References were ruled out if any of the above criteria were not met or if they did not report IRB/ethics committee approval and patient informed consent.<br>The primary outcome was device-oriented MACE at 12-month follow-up and the secondary outcomes were recurrent myocardial infarction (MI), probable or definite stent thrombosis (ST), and all-cause mortality (TD).<br>**Article: Study selection and Data extraction** |
| Information sources | 7 | *Describe all information sources (e.g., databases with dates of coverage, contact with study authors to identify additional studies) in the search and date last searched.*<br>Queries by FD & LL:<br>- MEDLINE, EMBASE, the Cochrane Library, Web of Science<br>- Also checked: AHA, TCT, ESC, EuroPCR, and clinicaltrials.gov<br>- Query on March 08 2018. Update query once all identified RCTs published on July 22 2020<br>- Contact with an investigator who participated in the majority of trials and as PI of some (PK)<br>**Article: Data sources.** |
| Search | 8 | *Describe methods used for assessing risk of bias of individual studies (including specification of whether this was done at the study or outcome level), and how this information is to be used in any data synthesis.*<br>Medline search on Pubmed:<br>((bioactive OR (Titanium AND nitride AND oxide) OR TiNO OR TNO OR BAS) AND stent) AND (DES OR (drug AND eluting AND stent)) AND (RCT OR ((randomized OR randomised) AND controlled AND trial)). No exclusion filter was applied related to |

| | | |
|---|---|---|
| | | language, country, year, or any other aspect.<br>The query was adapted to the search engines of the other 3 databases.<br>**Article: Data sources.** |
| Study selection | 9 | *State the process for selecting studies (i.e., screening, eligibility, included in systematic review, and, if applicable, included in the meta-analysis).*<br>Two reviewers (FD and LL) separately screened all references, classified them according to the review protocol's inclusion and exclusion criteria. Disagreements resolved by consensus after discussion with a 3rd reviewer (NM).<br>Selection helped with EndNote record classification functions. When duplicates identified, only one kept. When several references concerned the same study, their information was pooled using the most recent citation.<br>References ruled in: Study publications included if they met the following criteria: First-hand clinical evidence with prospective inclusion; patients with CAD treated with coronary PCI; implantation of either TiNOS or DES after random allocation; target outcomes reported at 1-year and/or 5-year follow-up; outcomes reported as the number of patients with an event or their rate along with the corresponding sample size.<br>References ruled out if any of the above criteria were not met or if they did not report IRB/ethics committee approval and patient informed consent.<br>All studies meeting PICOS-defined criteria included in the systematic review. Of those, all available data included in the meta-analysis.<br>**Article: Study selection and Data extraction** |
| Data collection process | 10 | *Describe method of data extraction from reports (e.g., piloted forms, independently, in duplicate) and any processes for obtaining and confirming data from investigators.*<br>Parallel, independent data entry by 2 reviewers (FD, LL) directly into RevMan 5.3 data tables. Then reconcilement of results and resolution of disagreements with third reviewer (NM). Evidence used was strictly that reported in publications. The similarity and compatibility of the definitions of endpoints across the eligible RCTs was confirmed with one investigator (PK).<br>**Article: Study selection and Data extraction** |
| Data items | 11 | *List and define all variables for which data were sought (e.g., PICOS, funding sources) and any assumptions and simplifications made.*<br>METHODS: Compliance of trial with PICOS specifications, coronary indication (ACS / other CAD), number of centers and countries. Date start/end of patient enrollment. Technical details of DES et TiNOS used. Description of endpoints; dates of follow-up visits and follow-up completion status. Statistical design, randomization and blinding methods. Trial registration & publication status.<br>BASELINE CHARACTERISTICS per treatment arm (DES vs. TiNOS): Number of patients randomized. Age, prior clinical events (prior MI, PCI or CABG), CAD clinical presentation (stable CAD, STEMI, NSTEMI, unstable angina, other CAD. Culprit lesion characteristics (RVD, LL, ACC/AHA lesion type), presence of thrombus, vessels treated. Procedure characteristics: number of stents per culprit lesion, TSL, procedural success, procedure success/failure, stent failure, DAPT duration and drugs, sample attrition and cross-overs.<br><br>OUTCOME VARIABLES per treatment arm (DES vs. TiNOS):<br>(a)Number of patients randomized (exposure = Intention to treat TT)<br>(b)Number of patients who received the treatment (exposure = per protocol PP)<br>(c)Number of patients followed-up at 1-year and 5-years (exposure = PP and available)<br>(d)Number of patients with each type of event (TLR-MACE, recurrent MI, probable or definite ST, cardiac death, TD) among those randomized to each treatment arm at each follow-up. |

| | | |
|---|---|---|
| | | The assumptions were that outcomes according to Academic Research Consortium definitions could be extracted.<br>Otherwise, that outcomes defined as they were in protocols were a reasonable proxy for outcomes defined per ARC.<br>- stratified data extraction according to two clinical indication subgroups: ACS vs. Other CAD<br>- copying the number of patients with an event for each outcome and each subgroup, or calculating those numbers from reported proportions in relation to the number of randomized patients.<br>- missing numbers could be obtained by subtracting the complement number from the total or adding up other numbers, and decimal numbers derived from rates could be rounded to the nearest integer.<br>It was also assumed that all studies would share would report TLR-MACE, recurrent MI, TD and ST according to similar definitions. The reviewers discussed this assumption with one investigator (PK).<br>**Article: Study selection and Data extraction** |
| Risk of bias in individual studies | 12 | *Describe methods used for assessing risk of bias of individual studies (including specification of whether this was done at the study or outcome level), and how this information is to be used in any data synthesis.*<br>Risk of bias in individual RCTs was assessed according to eight criteria proposed by the Cochrane Collaboration: (1) Random sequence generation (selection bias), (2) Allocation concealment (selection bias), (3) Blinding of outcome assessment (detection bias), (4) Incomplete outcome data (attrition bias), (5) Selective outcome reporting (reporting bias), (6) Performance bias was split into two components: (6a) blinding of operator versus (6b) blinding of participants and other personnel. The last item was (7) Other sources of bias. The split in performance bias was based on the fact that operators could not be blinded while other personnel and other caregivers could.<br>The risk of bias related to each item and for individual bias was entered directly in RevMan 5.3 using its piloted forms.<br>The cumulative risk of bias of individual studies was estimated with summary graphics. Items that caused the risk of bias were taken into account in when rating the level of certainty of overall evidence for each outcome when conducting GRADE analysis. Risk of bias was not quantitatively used to modify the relative weight of studies when pooling their results for each endpoint.<br>**Supplemental material : Part 1: Systematic review methods. Part 2: Figure 2a, Figure 2b.** |
| Summary measures | 13 | *State the principal summary measures (e.g., risk ratio, difference in means).*<br>The two treatment arms were compared for each endpoint using the risk ratio (RR) defined as *((n patients with an event in TiNOS)/(n patients in TiNOS))/((n patients with an event in DES)/(n patients in DES))*. Numbers were those that enabled an ITT analysis. The 95% CI of each RR was computed.<br>**Article : Statistical analysis.** |
| Synthesis of results | 14 | *Describe the methods of handling data and combining results of studies, if done, including measures of consistency (e.g., I2) for each meta-analysis*<br>Study RRs were pooled with the Mantel-Haenszel (M-H) fixed-effect model if there was no significant heterogeneity between the RRs of individual RCTs, and the M-H random-effects model otherwise.<br>The assumption was made that a fixed-effect model was applicable within ACS and within the “other CAD” subgroups respectively, but not when the two subgroups were pooled.<br>Heterogeneity between individual study RRs for each endpoint was tested using Cochran’s Q-test and the $I^2$ index (the percentage of heterogeneity between study RRs that is not attributable to random error). Heterogeneity was considered significant if $p < 0.05$ or if $I^2 > 50\%$.<br>**Supplemental material : Part 1: Systematic review methods.** |
| Risk of bias across studies | 15 | *Specify any assessment of risk of bias that may affect the cumulative evidence (e.g., publication bias, selective reporting within studies).*<br>Risk of bias across studies was rated for each endpoint separately during the GRADE assessment. (1)Cumulative risk of bias of |

| | | |
|---|---|---|
| | | individual studies estimated with summary graphics, (2) inconsistency with respect to the degree of heterogeneity between studies which may cause a bias in the pooled RR related to the predominance of some studies related to their relative weight, (3) indirectness between the evidence and the question to be addressed as defined according to PICOS. That risk concerned mainly MACE and recurrent MI whose definitions may have varied slightly across studies thus causing the studies with the largest relative weight to have an overrepresentation of their definitions , (4) Publication bias was estimated for each endpoint separately with a funnel plot and Harbord's test. An asymmetrical funnel plot and a Harbord regression test for dichotomous endpoints with p < 0.005 indicated a significant risk of publication bias<br>**Supplemental material : Part 1: Systematic review methods. Part 2: Figure 2a, Figure 2b.** |
| Additional analyses | 16 | *Describe methods of additional analyses (e.g., sensitivity or subgroup analyses, meta-regression), if done, indicating which were pre-specified*<br>Stratified/subgroup analysis and sensitivity analysis and were predefined in the protocol.<br>-All outcomes were analyzed with stratification on ACS and other CAD.<br>-The robustness of each outcome's pooled RR was assessed through sensitivity analysis, which consisted in iteratively recalculating the pooled RR after removing the input of one eligible RCT. As a result, the impact of each single RCT on the pooled RR of each outcome was quantified.<br>**Supplemental material : Part 1: Systematic review methods.** |
| **RESULTS** | | |
| Study selection | 17 | *Give numbers of studies screened, assessed for eligibility, and included in the review, with reasons for exclusions at each s tage, ideally with a flow diagram*<br>**Article: Figure 1 PRISMA flowchart.** |
| Study characteristics | 18 | *For each study, present characteristics for which data were extracted (e.g., study size, PICOS, follow-up period) and provide the citations.*<br>**Supplemental material : Table 1 Methods of the included RCTs** |
| Risk of bias within studies | 19 | *Present data on risk of bias of each study and, if available, any outcome level assessment (see item 12).*<br>**Supplemental material : Part 2: Figure 2a, Figure 2b, Table 3a GRADE assessment of certainty of evidence** |
| Results of individual studies | 20 | *For all outcomes considered (benefits or harms), present, for each study: (a) simple summary data for each intervention group (b) eff ect estimates and confidence intervals, ideally with a forest plot.*<br>**Article: Figure 2, Figure 3, Table 2. Supplemental material: Part 2: Figure 3a, Figure 3b, Table 3b GRADE summary of findings.** |
| Synthesis of results | 21 | *Present results of each meta-analysis done, including confidence intervals and measures of consistency.*<br>**Article: Figure 2, Figure 3, Table 2. Supplemental material: Part 2: Figure 3a, Figure 3b, Table 3b, GRADE summary of findings.** |
| Risk of bias across studies | 22 | *Present results of any assessment of risk of bias across studies (see Item 15).*<br>**Supplemental material : Part 2: Figure 2a, Figure 2b.** |
| Additional analysis | 23 | *Give results of additional analyses, if done (e.g., sensitivity or subgroup analyses, meta-regression [see Item 16]).*<br>**Article: Figure 2, Figure 3, Table 2. Supplemental material: Part 2: Figure 3a, Figure 3b** |
| **DISCUSSION** | | |
| Summary of evidence | 24 | *Summarize the main findings including the strength of evidence for each main outcome; consider their relevance to key groups (e.g., healthcare providers, users, and policy makers).* |

| | | |
|---|---|---|
| | | **Supplemental material : Part 2: Table 3a GRADE assessment of certainty of evidence, Table 3b GRADE summary of findings.** |
| Limitations | 25 | *Discuss limitations at study and outcome level (e.g., risk of bias), and at review-level (e.g., incomplete retrieval of identified research, reporting bias).*<br>**Article: Strengths and limitations of this study, Discussion. Supplemental material : Part 2: Table 3a GRADE assessment of certainty of evidence** |
| Conclusions | 26 | *Provide a general interpretation of the results in the context of other evidence, and implications for future research.*<br>**Article: Conclusion** |
| **FUNDING** | | |
| Funding | 27 | *Describe sources of funding for the systematic review and other support (e.g., supply of data); role of funders for the systematic review.*<br>This work was exclusively funded by the University of Bordeaux and INSERM BPH U1219 |

12
